# A CRISPR-Cas12a-based specific enhancer for more sensitive detection of SARS-CoV-2 infection

**DOI:** 10.1101/2020.06.02.20119735

**Authors:** Weiren Huang, Lei Yu, Donghua Wen, Dong Wei, Yangyang Sun, Huailong Zhao, Yu Ye, Wei Chen, Yongqiang Zhu, Lijun Wang, Li Wang, Wenjuan Wu, Qianqian Zhao, Yong Xu, Dayong Gu, Guohui Nie, Dongyi Zhu, Zhongliang Guo, Xiaoling Ma, Liman Niu, Yikun Huang, Yuchen Liu, Bo Peng, Renli Zhang, Xiuming Zhang, Dechang Li, Yang Liu, Guoliang Yang, Lanzheng Liu, Yunying Zhou, Yunshan Wang, Tieying Hou, Qiuping Gao, Wujiao Li, Shuo Chen, Xuejiao Hu, Mei Han, Huajun Zheng, Jianping Weng, Zhiming Cai, Xinxin Zhang, Fei Song, Guoping Zhao, Jin Wang

**Author notes:** Correspondence to: F. S.,; G.Z.,; J.W.,. [† These authors contributed equally to this work.].

## Abstract

High Ct-values falling in the grey zone are frequently encountered in SARS-CoV-2 detection by real-time reverse transcription PCR (rRT-PCR) and have brought urgent challenges in diagnosis of samples with low viral load. Based on the single-stranded DNA reporter *trans-cleavage* activity by Cas12a upon target DNA recognition, we create a <u>S</u>pecific <u>E</u>nhancer for detection of PCR-amplified <u>N</u>ucleic <u>A</u>cids (SENA) to confirm SARS-CoV-2 detection through specifically targeting its rRT-PCR amplicons. SENA is highly sensitive, with its limit of detection being at least 2 copies/reaction lower than that of the corresponding rRT-PCR, and highly specific, which identifies both false-negative and false-positive cases in clinic applications. SENA provides effective confirmation for nucleic acid amplification-based molecular diagnosis, and may immediately eliminate the uncertainty problems of rRT-PCR in SARS-CoV-2 clinic detection.

**One Sentence Summary:** CRISPR-Cas12a-based COVID-19 diagnosis.

## Main Text

Since December 2019, the outbreak of COVID-19, caused by the infection of the SARS-CoV-2, has rapidly spread throughout the world (*1*), and is now a global pandemic. Till June 1, 2020, the outbreak has affected 216 countries, areas and territories, infected 6 million people, and caused more than 370 thousand of death (*2*). One of the greatest public health concerns in combating against the pandemic is a prompt response to meet the urgent demand for rapid and accurate diagnosis of the virus. Currently, nucleic acid amplification-based molecular diagnostics (MDx) is the most accurate, fast and affordable and thus the preferred method for diagnosis of SARS-CoV-2 infection, and the real-time reverse transcription PCR (rRT-PCR) kits have been successfully developed by quite a few laboratories and commercial companies (*3*). However, since its clinical application at least five months ago in China, the diagnostic performance of rRT-PCR for SARS-CoV-2 has brought some urgent challenges, particularly with the frequently encountered high Ct-value designated “grey zone” associated uncertain negative or positive readouts (*4-8*). Besides of “human error” factors such as misconducted sampling, unqualified reagents and uncalibrated diagnostic equipment, inefficient RT reaction and PCR amplification of clinical samples with very low viral loads are likely the major intrinsic causative factors for the fuzzy rRT-PCR readouts and uncertain diagnosis. Although repetitive sampling and assays are implemented for final confirmation of the diagnosis, these “trouble shooting” efforts are time-consuming and may still fail to detect the low viral load samples from some mild or asymptomatic patients, or from the recovering patients, resulting in false-negative diagnosis that may cause serious public concerns in battling against the pandemic (**Figure 1**).

**Figure 1.**
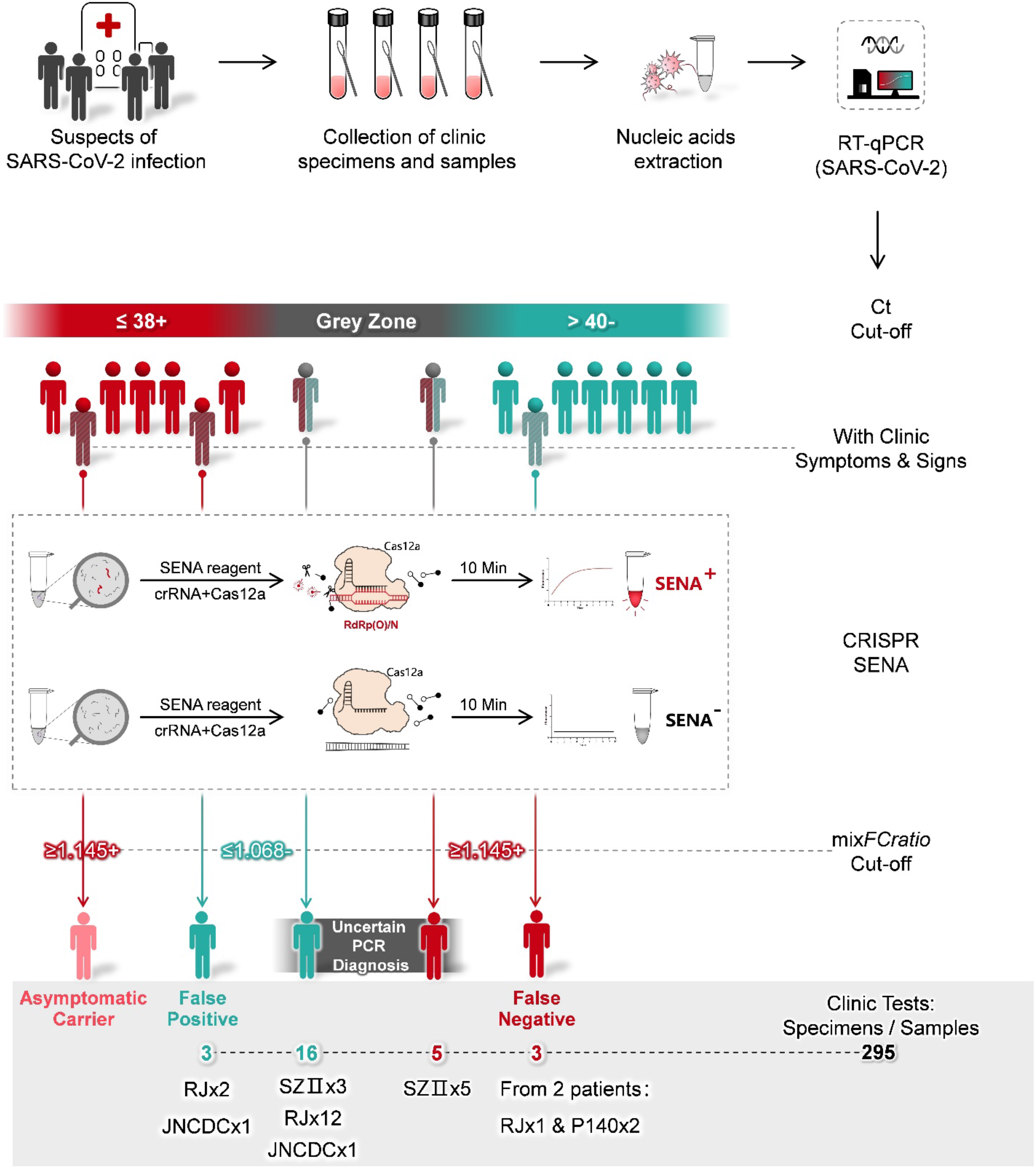
Schematic description of SENA and its application as a confirmation diagnosis for rRT-PCR diagnosis of COVID-19. Generally, nucleic acids are extracted from the clinic specimens such as pharyngeal swabs of the suspects of SARS-CoV-2 infection and then subject to rRT-PCR analysis. The diagnostic reports are based on the Ct cut-off values guided by the supplier of rRT-PCR kits. However, high Ct-value designated “grey zone” associated uncertain fuzzy readouts are often encountered. Besides, some probably false-positive or false-negative cases may be indicated by their atypical clinic symptoms or signs. For all these cases, the corresponding rRT-PCR products can be sent to another physically isolated room for SENA analysis and the ambiguity may be clarified by SENA with its positive and negative cut-off *mixFCratio*. The real-life data related to these scenarios revealed in this study are shown in the figure and details are illustrated in the text. RJ, JNCDC and SZII are the names of the hospitals and the number indicates the overall number of patients identified. While P140 was a patient in DF hospital, and two distinct samples from P140 were identified to be false-negative. For details, please ref to Supplementary **Table S3**.

With the characterization of non-specific *trans-cleavage* activities against single-stranded nucleic acids in several CRISPR-associated (Cas) proteins, *e.g*., Cas12, Cas13 and Cas14 (*9-15*), Clustered Regularly Interspaced Short Palindromic Repeats Diagnostics (CRISPR-Dx) technology (*16, 17*) was established and has been developing rapidly. The underline mechanism for CRISPR-Dx, as illustrated by the Cas12a-based HOLMES system for example (*18*), is based on the efficient trans-cleavage activity against a fluorophore quencher (FQ)-labeled single-stranded DNA reporter by Cas12a triggered upon target DNA recognition, which is guided by a specific CRISPR RNA (crRNA), generating exponentially increasing fluorescence signal within several minutes. With this mechanism, here, we design a <u>S</u>pecific <u>E</u>nhancer for detection of PCR-amplified <u>N</u>ucleic <u>A</u>cids (SENA) to improve both the detection sensitivity and specificity against the pre-amplified targeted SARS-CoV-2 genomic fragments (**Figure 1**). Briefly, the COVID-19 clinic samples are firstly analyzed by the well-established rRT-PCR assays and amplicons with uncertain readouts are then verified by SENA in a physically isolated space, avoiding contamination of the PCR laboratory during pipetting.

To prepare appropriate crRNAs for SENA detection, we firstly determined the amplicon sequences from several commercial rRT-PCR kits used in China and then designed specific crRNAs corresponding to each of the distinct amplicons (**Table S1**). Candidate crRNAs were prepared and analyzed in a SENA system comprised of Cas12a, crRNAs, FQ-reporter and the rRT-PCR products using templates of either the positive or negative controls, and the best crRNAs, *i.e*., the lowest fluorescence with the negative control and highest with the positive control, were chosen for further SENA assays (data not shown). If a kit detects more than one gene, *e.g*., Orf1ab (“abbreviated as *O*”), *E* and *N* genes, corresponding crRNAs were mixed in a SENA system to enhance the detection signal.

The performance of SENA was quantitatively characterized *via* a systematic titration upon rRT-PCR amplicons employing pure SARS-CoV-2 RNA standards comprised of the *O* and *N* fragments as the templates. As it is aware that the viral nucleic acids extracted from patients’ samples such as nasopharyngeal swabs usually contain some biological and chemical contaminants that might inhibit the enzyme activities for reverse transcription and PCR reactions (*19*) and is likely one of the causal effects attributed to the low efficiency of rRT-PCR in clinical analysis. In order to mimic the clinical sampling for the titration experimentation, the RNA standards were serially diluted in buffer prepared by mixing the nucleic acid extracts from 40 COVID-19 negative people, generating RNA templates ranging from 0.025 to 25 copies per reaction (Rx).

Due to the Poisson distribution property of sampling, replica variations become extremely significant when the template copies in individual reaction are designed to be low, *i.e*., less than 3-4 copies/Rx, near the limit of detection (LoD) for rRT-PCR (20, 21), and extremely low, *i.e*., equal to and less than 1 copy/Rx. To overcome this sampling ambiguity problem, we performed 9 replicas for groups with 1 and 0.5 RNA template copies/Rx while 6 replicas for each of the rest concentrations. In addition, although the rRT-PCR assay supplier, Shanghai BioGerm (BJ), who follows the Chinese CDC recommended primer sets (**Table S1**), recommends 40 cycles of PCR amplification, we set 45 cycles as routine aiming at recording maximum exact Ct values if possible. After rRT-PCR reaction, all amplicons were subjected to 3 individual SENA reactions, *i.e*., N-SENA, O-SENA and mix-SENA with crRNAs targeting *O* gene, *N* gene and both, respectively.

Consistent with the theoretical analysis (*20*) and rigorous experimentation (*21*), along with the decrease of the RNA templates to less than 3 copies/Rx, the rRT-PCR Ct values in some replicas, primarily that corresponding to the *N* gene, passed 38 (the cut-off for positive as recommended by the rRT-PCR kit suppliers) but were less than 40, which should be considered as entering the “grey zone”. The Ct values increased steadily when the concentration of the RNA templates further decreased, with more and more replicas showing one or both Ct values entering the “grey zone” and eventually all became “negative”, *i.e*., greater than 40 or even 45 (**Figure 2A, Table S2 and Figure S1**). Employing Ct=38 as the cut-off for “positive” detection, we estimated the LoD for *O* and *N* genes with 95% confidence interval (CI) of this set of rRT-PCR assay as 3.3≤4.0≤6.1 and 4.0 ≤ 4.1 ≤ 4.4, respectively (**Figure S2**). Most likely due to the influence of the complex combination of the targeted viral genomic fragments and the clinic sampling background, the LoD determined in this study was clearly higher than the published value of 2.0≤2.5≤3.7, which analyzed single target in a pure system (*21*).

**Figure 2.**
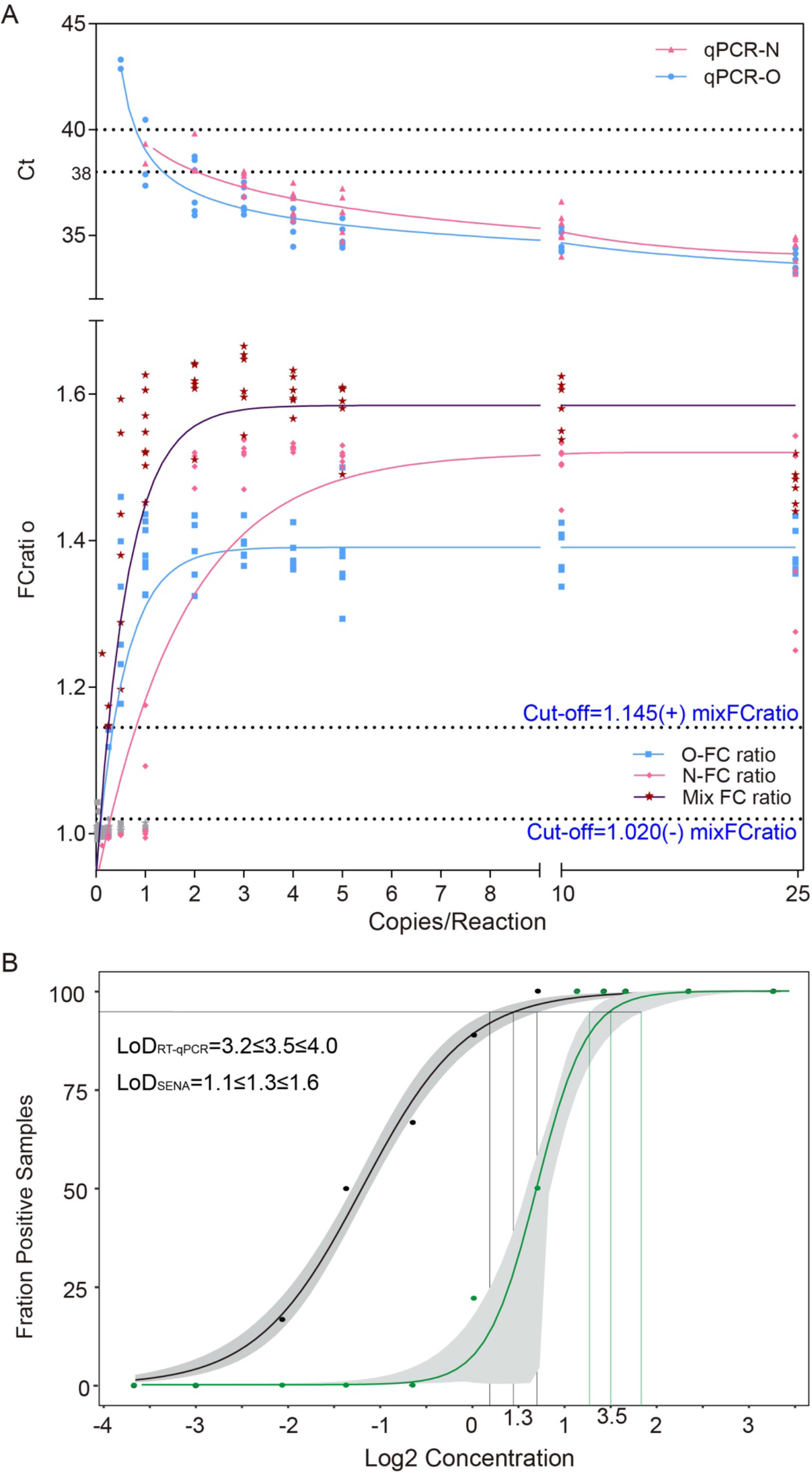
Determination of the cut-off values for SENA detection (A) and the LoD values with 95% CI for both rRT-PCR (O-Ct, green dots) and SENA (mix-FCrafio, black dots) (B) based on the systematic titration assays. All the experimental and analytical details are described in the text. Notice that the SENA negative cut-off was set as mix-FCratio=1.020 in this figure on the basis of the titration assay of the standard RNA templates but was adjusted to 1.068 along with the increase of the clinic applications (**Figures 1** and **4**, Supplementary **Table S3**).

The rRT-PCR amplicons were further analyzed by SENA detection with the measurement of the fluorescence signals for each corresponding replica. After comparison of the parameters of slope (increase of fluorescence/min) versus FC (the fold of change of fluorescence between that of the sample over that of the negative control at certain time point), we defined a parameter, *FCratio*, which is the ratio of the FC at 10 min to that at 5 min after the initiation of fluorescence reading (**Figure S1** and **Table S2**). We also found that in the cases with low concentrations of templates, the rRT-PCR efficiency of the two target genes (*i.e., O* and *N*) were different so as the SENA detection (**Figures S1 and S2**). In order to verify the existence of specific amplicons of SARS-CoV-2 nucleic acids in an individual rRT-PCR reaction, all of the amplicons of the replicas with RNA templates ranging from 0.125 to 2 copies/Rx were subjected to next generation sequencing (NGS) analysis. The results were found to be completely consistent with the perspective results of both O-SENA and mix-SENA. In addition, with mix-SENA, not only the signals are generally more significant than that of the O-SENA detection but also may resolve some of the ambiguity readouts found with N-SENA (Figure S1 and Table S2). Based on these results, the *mixFCratio* was demonstrated as the most sure-proof index for rRT-PCR confirmation, and we empirically estimated that mixFCratio≥1.145 for positive cut-off, and *mixFCratio≤1.020* for negative cut-off (**Figure 2A**). Of course, these two parameters are subject to further verification and adjustment along with the increase of tested samples. Because SENA is rRT-PCR based, the same methodology for determining the rRT-PCR LoD was used to estimate that of SENA by this set of data, corresponding to both individual *O* and *N* fragments (**Figure S2**) and in combination as indicated by the mix-SENA (**Figure 2B**). As expected, the N-SENA LoD (3.7≤4.3≤4.8 with 95% CI) is very close to that of the N-Ct of rRT-PCR, while the LoD of O-SENA (1.1 ≤ 1.3 ≤ 1.7 with 95% CI) is significantly lower than that of O-Ct (**Figure S2**). Although the LoD of mix-SENA (1.2≤1.6≤2.1 with 95% CI) is slightly higher than that of O-SENA (**Figure 2B** and **Figure S2**), it is apparently caused by its capable of confirming some of the ambiguous amplicons in the extremely low concentration cases (**Figure S2**) and thus, *mixFCratio* is chosen for clinic applications.

SENA was further verified in a few hospitals, testing various clinic specimens and samples under different scenarios (**Figure 1**) and employing few more commercial rRT-PCR diagnosis kits in addition to BJ which was used in the titration experiment (**Table S1**). Totally 295 clinic samples or specimens (mainly pharyngeal swabs) collected from 282 individuals were tested by rRT-PCR followed by SENA detection (**Table S3**). Except for asymptomatic carriers, all the cases of uncertain analytic and false positive or negative readouts of rRT-PCR diagnosis were encountered and finally confirmed or corrected by SENA detection.

Specifically, samples from 139 patients of Ruijin Hospital (RJ, Shanghai, China) were assayed by rRT-PCR employing diagnostic kits of ZJ and HD, 137 of which had consistent readouts by both rRT-PCR kits, indicating two positive, 123 negative and 12 suspected that fell in the “grey zone” (**Table S3**). SENA detection of these samples revealed not only the 12 suspected as negative but also identified one more positive among the original 123 negative individuals, clearly a case of false negative diagnosis (**Table S3**). Besides, distinct rRT-PCR assay results, positive by HD but negative by ZJ were shown for samples collected from 2 close contacts of COVID-19 patients and apparently asymptomatic (ref to **Table S3**). However, the amplicons of both ZJ and HD were shown as negative via SENA detection. All these ambiguous rRT-PCR amplicons (17 samples, ref to **Table S3**) were finally analyzed by NGS, and the results were consistent with the SENA. Noticeably, the rRT-PCR false-negative COVID-19 patient was symptomatically mild at the point of admission with all the clinic laboratory tests negative but turned positive after 24 hours. On the other hand, although those 12 suspected patients had respiratory infection symptoms, they were finally excluded from COVID-19 according to the latest guideline for diagnosis and treatment from China National Health commission (the 6th edition). Similarly, in Shenzhen Second People’s Hospital (SZII, Shenzhen, China), 5 uncertain rRT-PCR readouts for *O* gene were found among 139 individuals. Three of them had Ct value of 39.47, 39.7 and 40.56, respectively but the following SENA detection gave *mixFCratio* values less than 1.0 for all of them, indicating all negative. The other two individuals had Ct values of 38.87 and 39.22, while their *mixFCratio* values were 1.581 and 1,609, respectively, indicating positive for both. In addition, there were another three individuals with Ct values larger than 40 for *O* gene and 36.09, 35.88 and 37.98 for *N* gene, respectively; however, the following SENA detection showed *mixFCratio* values were 1.39, 1.55 and 1.21, respectively, indicating all positive. All these amplicons were further confirmed by NGS analysis (**Table S3**), obtaining consistent results with those of SENA. Consistently, the three SENA-negative individuals were finally excluded from SARS-CoV-2 infection after being rechecked by rRT-PCR after 24 hours (**Table S3**). Based on above data, it is clear, SARS-CoV-2 infection suspects with either rRT-PCR Ct values falling in the “grey zone” or with clear patient-contact epidemiological history but negative rRT-PCR tests, are strongly recommended to perform SENA detection to minimize the possibility of misdiagnosis. On the other hand, in case an rRT-PCR-positive suspect does not demonstrate any COVID-19 clinic symptoms and/or signs, SENA detection is also strongly recommended to eliminate either false-positive diagnosis or misdiagnosis of the so-called “asymptomatic carrier” or “asymptomatic patient”.

Besides of preventing false-negative or false-positive diagnosis, the highly sensitive property of SENA may also assist in providing evidence of viral clearance for COVID-19 recovering patients. A female patient in Dongfang Hospital (DF, Shanghai, China) was confirmed as COVID-19 positive by both rRT-PCR and CT scanning and showed ground-glass opacities mixed with consolidation along the subpleural area (**Figure 3**). Accordingly, the SENA test was positive with the *mixFCratio* of 1.43. After the hospitalization, the patient was further analyzed by rRT-PCR at two time points, obtaining all negative results with bilateralnasal and pharyngeal swab specimens. However, the *mixFCratios* of SENA for some of her specimens were 1.64, 1.36 and 1.00, respectively, indicating that the virus was contained and yet to be cleared. On the sixth day, both rRT-PCR and the corresponding SENA detection for all of her specimens were negative and these results were confirmed by NGS and consistent with her normal CT scanning results (**Figure 3**). Thus, she was discharged from the hospital and safely back to home. Similar cases were found in Jinan of Shandong Province, China, where the fecal samples from two recovering COVID-19 patients were tested negative by rRT-PCR but clearly positive by SENA (**Table S3**). Considering a certain percentage of the recovered patients discharged from hospitals were reported to be re-detectable positive (RP) (*22*), the incomplete clearance of the SARS-CoV-2 virus ahead of discharge might be one of the possible causes. Therefore, it could be necessary to consider more sensitive detection approaches such as SENA as a potential index of viral clearance.

**Figure 3.**
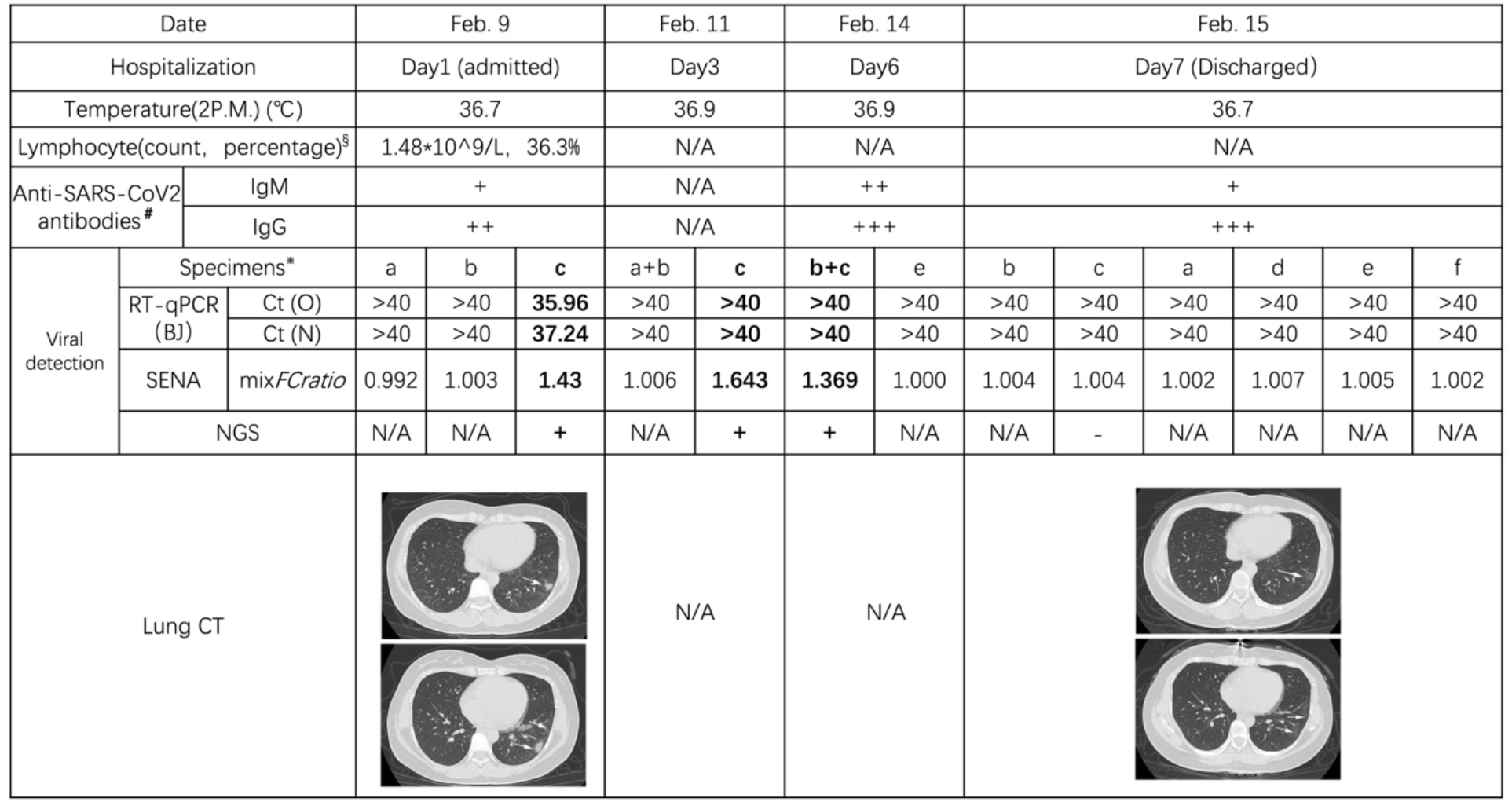
Schematic diagram of the hospitalization process of patient P140 (Shanghai DF Hospital) Detailed viral detection data are listed in **Table S3**. Other clinic data indicated that P140 is a COVID-19 patient with mild clinic symptoms. ^§^Reference range for Lymphocyte count: 1.1-3.2×10^9/L; Reference range for the percentage of Lymphocyte: 20-50%. ^#^Label of the antibodies: +, weak; ++, medium; +++, strong. ^※^Label of the specimens: a, pharyngeal swab; b, nasal (left) swab; c, nasal (right) swab; d, serum; e, plasma; f, fecal.

To reconfirm and/or improve the cut-off values for SENA *mixFCratio*, the ambiguous Ct values were re-estimated using the regression functions derived from the rRT-PCR assays with titrated standard RNA templates (**Figures S4** and **S5**), and then the Ct values (both estimated and detected) were plotted against the corresponding *mixFCratios* (**Figure 4**). Combining the data from both RNA standards and clinic samples, it is clear that SENA detection is of both high sensitivity, identifying real positive samples with Ct values as high as more than 43 (approaching 50 as estimated), and high specificity, identifying real negative samples with Ct values as low as 39. Therefore, SENA can effectively eliminate uncertain diagnosis of rRT-PCR assays for SARS-CoV-2 infection. In addition, the cut-off value for SENA *mixFCratio* remains unchanged as 1.145 for positive diagnosis while slightly increased to 1.068 for negative (**Figure 4**), which is supposed to further increase along with the clinic applications.

**Figure 4.**
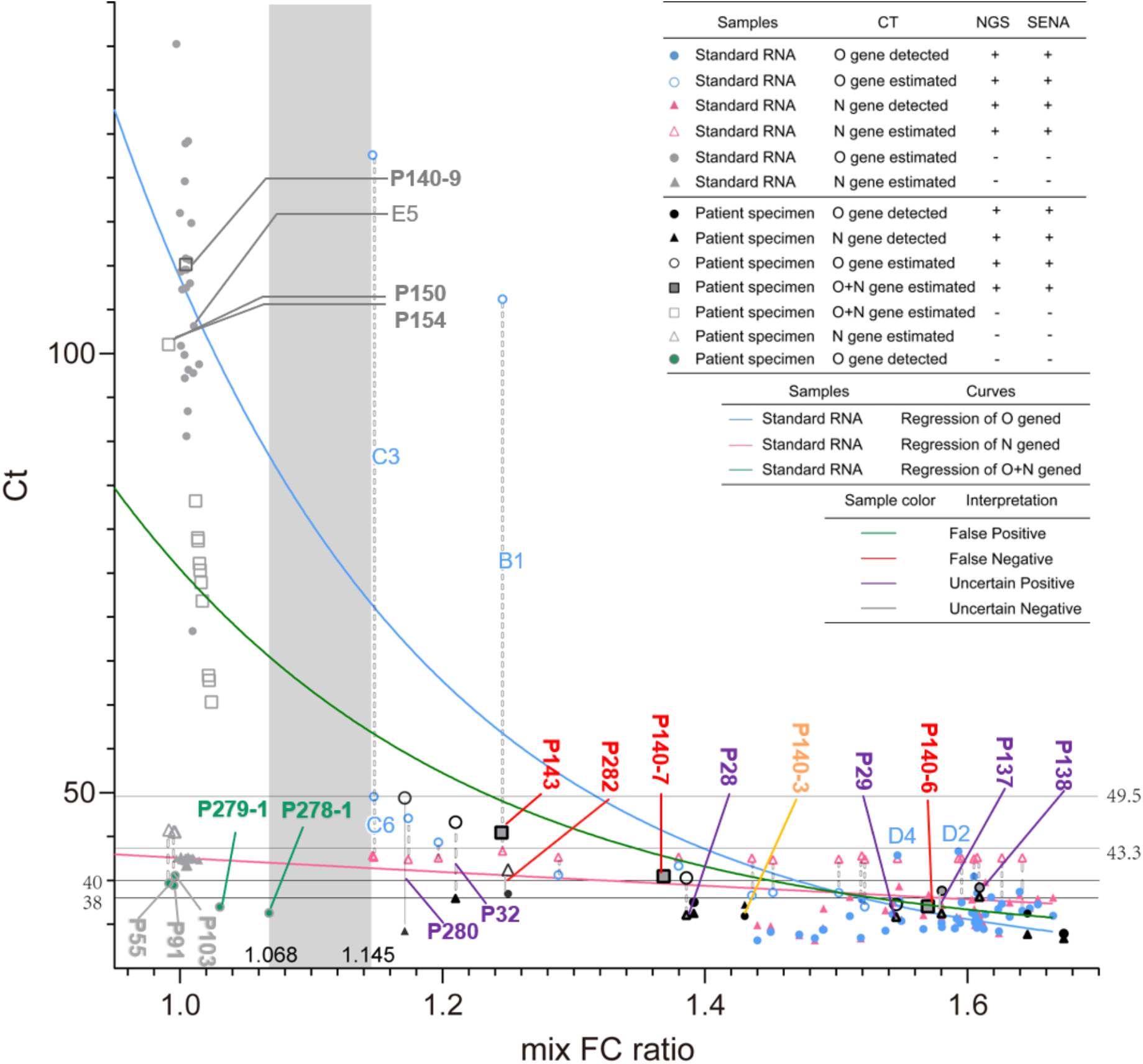
Apparent correlation plot of the rRT-PCR Ct values against the SENA *mixFCratios* in SARS-CoV-2 detection. All the data of the systematic titration experiment with low concentrations of standard RNA templates (**Table S2A**) and the data of clinic tests employing samples with ambiguous rRT-PCR readouts are used in this plot. In case the Ct values are too high to be detected by the rRT-PCR assay, i.e., Ct>40~45, depending on the scenarios, the *mixFCratio-* correlated “apparent Ct values” may be estimated *via* the template concentration-related regression functions (**Materials and Methods 4.5**); however, majority of the extremely high “apparent Ct values” in real negative samples are arbitrary and are adopted merely to simplify the presentation. The positive cut-off of the mix-SENA detection (mixFCratio=1.145) is defined by the C3 and C6 samples of the systematic titration experiment (**Figure 2A**), while the negative cut-off of the mix-SENA detection (mixFCratio=1.068) is defined by the P278-1 sample of the clinic tests (**Table S3**). This plot confirmed the cut-off Ct values of rRT-PCR test provided by the kit supplier (BJ, ref to all detectable Ct values shown as solid dots). Meanwhile, with the aid of mix-SENA, the sensitivity of rRT-PCR was increased up to the detected level of O-Ct=43.3 in samples of D2 and D4 and estimated level of O-Ct=49.5 in samples of C6 and P280. In addition, false positives were detected with O-Ct values as low as 39 (P278-1), which was also the key test to define the negative cut-off of mix-SENA.

Instead of developing a closed CRISPR-Dx system, which, ideally, should be comprised of both target nucleic acids amplification and CRISPR-Cas-based *trans*-cleavage assays, SENA was created here to match the commercially available and widely applied rRT-PCR kits, and to solve the uncertainty challenge of the rRT-PCR “grey zone” in COVID-19 diagnosis. Although there are dozens of commercial rRT-PCR kits using distinct PCR primers and probes, their corresponding SENA detection kits can be easily designed and prepared following the instructions provided in this study. Considering the fact that qPCR is the most popular MDx system and SENA is simple to operate, SENA thus has the potential to be widely used in various scenarios to solve the uncertainty problems of qPCR as well as other nucleic acid amplification-based MDx. Of course, to minimize the possibility of aerosol contamination during opening of the PCR tubes and pipetting, physical separation of the SENA detection center from the clinical PCR laboratory is an absolute requisition. Abided by this rule, SENA has been demonstrated convenient and effective in several hospitals and centers for disease control and prevention as part of their laboratory routine in combination with rRT-PCR for more sensitive and accurate detection of SARS-CoV-2 infection. Therefore, SENA is a useful technique that meets the urgent needs of combating COVID-19 pandemic.

## Data Availability

All data is available in the main text or the supplementary materials.

## Acknowledgments

We thank Yucai Wang and Linxian Li for discussions and support. J.W. is supported by the National Natural Science Foundation of China (31922046); W.H. is supported by the National Key R&D Program of China (2019YFA0906000) and NSFC (81772737); D.W. is supported by NSFC (81972830) and Shanghai “Rising Stars of Medical Talent” Youth Development Program-Clinical Laboratory Practitioners Program; F.S. is supported by NSFC (31670757); G.Z. is supported by the Strategic Priority Research Program of the Chinese Academy of Sciences (XDB19040200).

## Data and materials availability

“All data is available in the main text or the supplementary materials.”

## Supplementary Materials for

**This PDF file includes:**

Ethics statement

Materials and Methods

Figs. S1 to S5

Tables S1 to S3

### Ethics statement

The human pharyngeal swab samples were collected from residual samples that had been obtained for clinical respiratory virus detection. All procedures complied with the Measures for the Ethical Review of Biomedical Research Involving Human Subjects issued by the National Health and Family Planning Commission of The People’s Republic of China. All protocols were approved by the Ethics Committees of Ruijin Hospital, Dongfang Hospital and Shenzhen Second People’s Hospital. All procedures were performed in biosafety level 2 facilities.

## Materials and Methods

### 1. RT-qPCR reactions

Reactions were conducted in a 25-μl reaction mixture following the instructions offered by the commercial suppliers of the reaction kits (ref to Supplementary **Table S1**). Usually, the reaction cycle parameters were set as reverse transcription at 50 °C for 10 min, denaturation at 95 °C for 5 min, then followed by 45 cycles of amplification, *i.e*., 95 °C for 10 s and 55 °C for 40 s.

### 2. SENA reagents and detection

#### 2.1 Preparation of SENA reaction reagents

Candidate crRNA guide sequences with the “TTN” PAM sequences were designed as shown in Supplementary **Table S1**, and the crRNAs were prepared following the procedures previously described (*11*). Except the target DNA, the 2× SENA reagent comprises of 2× NEB buffer 3.1, 500 nM LbCas12a (Tolo Biotech.), 1 μM synthesized crRNAs (for each specified target and thus varies according to the RT-qPCR kit supplier, **Table S1**), 1 μM FQ-reporter, and 1 U/μl RNase inhibitor (TaKaRa).

#### 2.2 SENA detection

To avoid the aerosol contamination of the MDx laboratory, after the RT-qPCR reaction, their products must be transferred to a physically isolated room to perform SENA detection. It is also important to choose proper SENA detection reagents corresponding to the RT-qPCR kits. To prepare a 20-μl SENA reaction system, with corresponding positive and negative controls, 2-μl PCR products and 8-μl RNase-free H_2_O were mixed with 10-μl 2× SENA reagent, and the mixture was then measured on an appropriate fluorescence reader with FAM fluorescence collected following the programs: 48 °C 30 s per cycle, 20 cycles. Both the slope and the Fluorescence Change (FC) can be calculated at any time points as desired.

### 3. Next generation sequencing (NGS)

The RT-qPCR products were purified by AMPure XP beads (Beckman Coulter Life Sciences, US), and libraries were then constructed following the procedures of end repair, dA-tailing and adaptor ligation, with the StepWise DNA Lib Prep Kit for Illumina (ABclonal, China). After PCR amplification, samples were sequenced on Illumina Miniseq to produce 2×150 bp paired-end reads. After adaptor trimming and quality trimming, the clean reads were mapped to the reference genome of SARS-CoV-2 (MN908947.3) using Bowtie2 (*22*).

### 4. Systematic titration and regression analyses

#### 4.1 Systematic titration experimentation

##### 4.1.1 The standard RNA templates

The SARS-CoV-2 RNA standards were purchased from Genewell (Shenzhen, China). According to the supplier’s information, three plasmids containing the whole sequences of *N* and *E* genes, and partial sequence of the Orf1ab, i.e., from 13237 to 13737 of the SARS-CoV-2 complete genome (MN908947.3), were transcribed *in vitro* individually. The RNA products were mixed with equal molar, aliquoted with addition of 1 μg of human RNA per tube, and subject to lyophilization and subsequent quantification with digital PCR. This SARS-CoV-2 RNA standard dry powder containing 1808 copies of *O* gene, 179.5 copies of *N* gene and 1160 copies of *E* gene was dissolved with 10 μl RNase-free water to obtain the original stock solution (estimated 180.8 copies/μl of O, 179.5 copies/μl of *N* and 116 copies/μl of E).

##### 4.1.2 The preparation of the serially diluted RNA templates

Pharyngeal swab samples were collected from 40 adult patients in Shenzhen Second Peoples’ Hospital by the Clinic Diagnosis Laboratory and the nucleic acids of each sample were extracted with the pre-packaged nucleic acid extraction kit (Da’An Gene., Ltd., Guangzhou, China), according to the manufacturer’s instructions, ended up with 55-μl extracts per sample. After RT-qPCR assays employing 5 μl of the extracts from each sample, all of the samples were shown to be SARS-CoV-2 negative. The remaining 50-μl extracts of each sample were mixed together and 5 μl of the mixture was once again analyzed by RT-qPCR and confirmed to be SARS-CoV-2 negative. Then, the mixed nucleic acid extract was used as the dilution buffer (totally about 2 ml) for serial dilution of the SARS-CoV-2 RNA standard stocks, generating desired concentrations (i.e., 5, 2, 1, 0.8, 0.6, 0.4, 0.2, 0.1, 0.05, 0.025, 0.01, 0.005 copies/μl), and 5 μl of each of the diluted solutions were used as templates for RT-qPCR analysis, forming gradient template concentrations (i.e., 25, 10, 5, 4, 3, 2, 1, 0.5, 0.25, 0.125, 0.05, 0.025 copies/Rx).

##### 4.1.3 Replica setting

We analyzed 9 replicas for each of the concentrations of 1 and 0.5 RNA template copies/Rx while 6 replicas for each of the rest concentrations (Supplementary **Figure S1**).

##### 4.1.4 RT-qPCR reactions

RT-qPCR reaction kit was supplied by BJ (**Table S1**), who follows the primer sets recommended by Chinese CDC. Instead of the recommended 40 cycles of PCR amplification, we set 45 cycles as routine aiming at recording the maximum exact Ct values if possible.

##### 4.1.5 SENA detection

Every amplicon of the RT-qPCR reactions was subjected to SENA detection. Specially, for this experimentation, three sets of SENA reagents were individually used, O-SENA contains the crRNA targeting the *O* sequence, N-SENA contains the N-targeting crRNA, and the mix-SENA contains crRNAs for both sequences.

#### 4.2 Choice of *FCratio* as the standard readout for SENA detection and mix-SENA as the standard reagent for clinic application

Three readout parameters were compared as shown in **Figure S1** (original data in **Table S2**). The slope (increase of fluorescence/min) represents the reaction rate of Cas12a *trans-cleavage* activity, but in this experiment, it represents neither the initial rate of the enzyme under limited substrate condition nor the pure first-order reaction rate varies according to the substrate concentration, particularly, demonstrated in cases of high template concentration, in which, the slope goes down along with the increase of the template concentration. In addition, when the substrate concentration is low, the slope of SENA is hard to be distinguished from that of the negative control, ending with ambiguous cut-offs. The FC (the fold of change of fluorescence between that of the sample over that of the negative control at certain time point, usually 5-30 min) does show clear differences between the positive amplicons from that of negative control, and it also shows certain quantitation character particularly at the low concentration templates cases. Because of these properties, FC has been a parameter used by a few users of CRISPR-Dx (*24*). However, it seems that the absolute value of the FC usually varies along the reaction time and sometimes it is influenced by the change of the fluorescence signal of the negative control. Besides, it is difficult to determine the “best choice” of the FC recorded at certain time points, which may cause confusing in clinic applications. It is clear, we need a stable readout which reflects the dynamic process and the quantitative correlation of SENA reaction with the low concentration of the templates on one hand and should be robust and accurate for clinical diagnosis on the other hand. We defined *FCratio*, which is the ratio between FC’s of SENA detection at 10 min versus 5 min after the beginning of the fluorescence reading. It not only measures the fluorescence change of SENA against the negative control background so that the quantity of the amplicons, particularly at the low concentration range may be represented, but also normalizes the slope of the fluorescence change of SENA to eliminate the complex background differences. As shown in Figure S2, *FCratio* significantly amplified the positive signals and represents the quantitation of the amplicons at low template concentrations to certain extent.

As shown in **Figure S1**, the capacity of the three SENA reactions are compared. It is obvious that N-SENA is the least sensitive one, while although O-SENA seems much more sensitive than that of N-SENA and largely comparable to that of mix-SENA, its signal at the very low template concentration range seems uncertain in some cases. Therefore, for clinic application, mix-SENA is the choice.

#### 4.3 Quality analyses of the RT-qPCR titration data

The quality of the Ct values vs the concentration of the standard templates for RT-qPCR of the systematic titration experiment were analyzed both empirically and statistically. Firstly, as shown in **Table S2** and **Figure S1**, the amplification efficiencies of the two genes represented by the valuable Ct readouts were different. The apparently lower sensitivity of the *N* gene amplification is in contrast with the clinic experiences and might due to the difference in the property of the templates used in different experiments (laboratory standard RNA template vs clinical real viral template). Secondly, although linear regression can be readily made between the Ct-values and the log10 (conc) (**Figure S3A**) as that of the previously published tests, the quality of the regression as judged by the R^2^s (**Figure S3A**) and the residues (**Figure S3B**) are clearly suboptimal likely due to the limited number of replicas in the experimentation. On the other hand, this titration was designed with taking at least the two most fundamental limitation factors about the sensitivity of COVID-19 RT-qPCR diagnosis into consideration, *i.e*., the sampling ambiguity and the influence of the biological/chemical contaminants from the clinic samples. Therefore, the data will be used for determination of the LoD for RT-qPCR and SENA (**Materials and Methods 4.4 and Figure S2**), and the regression function will be used to estimate the “apparent Ct” values of the samples with Ct-values greater than 45 (no amplification signal) but their SENA detection is positive (**Materials and Methods 4.5 and Figure S4**).

#### 4.4 Determination of limit of detection (LoD) for RT-qPCR and SENA

The LoD values for RT-qPCR (N-Ct and O-Ct) and SENA (N-*FCratio*, O-*FCratio* and *mix-FCratio*) were estimated based on the systematic titration employing standard RNA templates (**Figure S1 and Table S2**). The fractions of positive replicates versus the number of target molecules (copies) per reaction for N and O gene of COVID-19 were plotted and used the **sigmoidal function (1)** to fit the data via R (software version 3.5.0). The 95% confidence intervals were derived by bootstrapping the model residues and were visualized by R (software version 3.5.0) with built-in ggplot2 library (*21*).

#### 4.5 Regression of RT-qPCR Ct-values and the SENA *FCratio* versus the concentration of the templates employing the data from the systematic titration

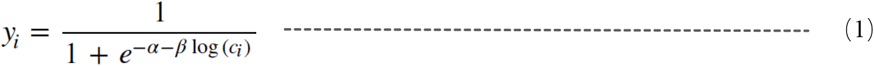

#### 4.5.1 Regression of RT-qPCR Ct-values with the concentration of the templates (copies/Rx)

Since PCR product increased exponentially with the initial concentration of the sample (x), and the Ct value of RT-qPCR parameter (y) was inversely correlated with the initial concentration, especially in the range of low copy number (low template concentration) samples, the power function equation (a<1) should be suitable for the data fitting. However, some of the experimental groups included very low initial sample concentrations (<1 copy/Rx), those amplification efficiencies should be different (particularly affected by sampling ambiguity) from that of the groups with high initial template concentration. Therefore, the power function formula with four parameters (Y=aX^n^+bX^m^) was used to match all the experimental group data to obtain a more accurate data model (**Functions 1, Figure S4**).

##### 4.5.2 Regression of SENA *FCratio* with the concentration of the templates (copies/Rx)

The exponential function (first order association kinetics of the interaction between a substrate and an enzyme, Y=a+b(1-e^cX^)) is used to fit the data of *FCratio* against the concentration of the templates. At low concentration (especially when the concentration is less than 2 copies/Rx), the *FCratio* is positively correlated with the template concentrations. However, when the template concentration reaches to 2 copies/Rx and more, the *FCratio* does not increase accordingly and the curve tend to be flatted out. In addition, as *FCratio* have been already normalized by the fluorescence signal of the negative background, it is stable and, in this case, we give the parameter Y_0_ being set as a constant value between 0.9 to 1 (**Functions 2, Figure S5**).

##### 4.5.3 Regression of RT-qPCR Ct-values with the SENA mix*-FCratio* values

In practice, quite significant portions of the clinic positive samples detected by mix-SENA with their *FCratio* readings higher than the positive cut-off, but with a negative PCR Ct value (>40-45, depending on the scenario). Under certain circumstances, people may be interested to learn the copy number of the templates for the corresponding RT-qPCR assays or even the “probable” Ct values of these assays. With the aid of the above-mentioned two regression functions (1 in **Figure S4** and 2 in **Figure S5**), these data could be estimated. One may firstly substitute the Y in Function 2 by the measured *FCratio* value and the corresponding X can be calculated representing the “estimated concentration of the template). Then this X value can be used to estimate the corresponding “estimated Ct-value” as the Y of Function 1. We estimated all the ambiguous Ct-values of the positive amplicons and plotted them against their corresponding mix-*FCratio* (**Figure 4**). All the real and estimated Ct values for both *N* and *O* genes are plotted against the corresponding *FCratio* values of mix-SENA as X axis. An exponential decay function (with X_0_ = 1; When X≤X0, Ct=∞; otherwise, one phase decay) fits well to all the data (R^2^=0.9238) and is used for analyzing the clinic data and adjust the cut-off values accordingly (**Figure 4**).

#### 5. Detection of antibodies

The detection of anti-SARS-CoV2 antibodies was executed by the point-of-care microfluidic platform integrating a home-made fluorescence detection analyzer (Suxin, Shanghai, China). A total of 10-μl plasma was added into the loading chamber of microchip followed by the addition of 70-μl sample dilution buffer. After incubation for 15 min at room temperature, the microchips were loaded onto the fluorescence detection analyzer, and fluorescence signal was detected from the analyzer, following the manufacturer’s instruction.

## Supplementary Figures

**Fig. S1.**
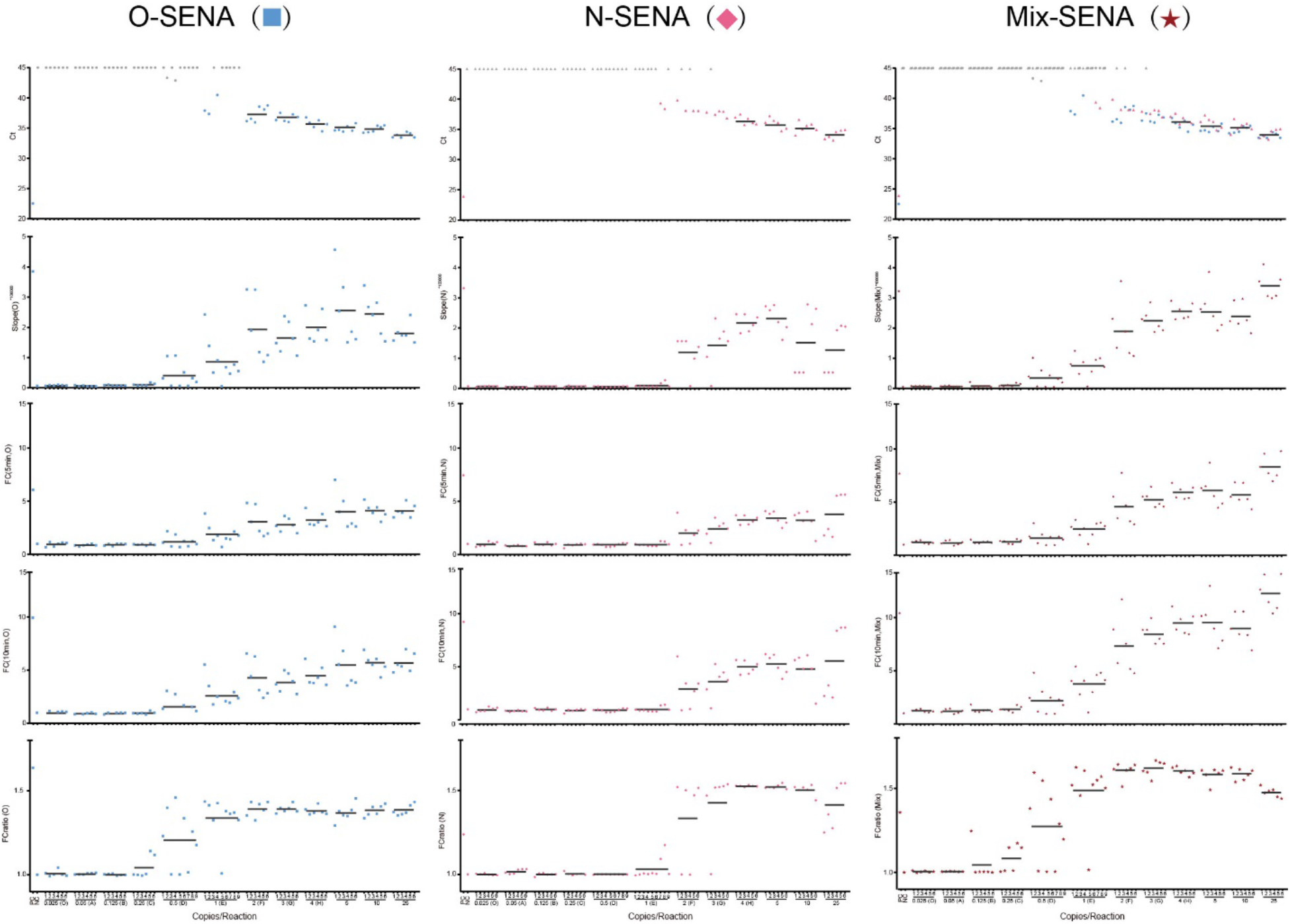
Systematic titration of serially diluted SARS-CoV-2 standard RNA templates detected by RT-qPCR followed by SENA. The design of this experiment is illustrated in detail in *Materials and Methods 4*. All the resulted readouts are listed in Table S2 and are plotted in this figure for individual tests being numbered with each of the concentration of the templates used as marked in the bottom X-axis. The very top horizontal row of the panel matrix is the test of RT-qPCR with Ct values detected via fluorescence of FAM (● for ○ gene), VIC (▲ for N gene) and both, respectively. In case Ct≥ 45, *i.e*., no fluorescence signal of PCR amplification could be detected, the corresponding symbols were shown in grey (**●** or **▲**). The vertical lists from the second row of the panel matrix represent the SENA reagent used for the detection: O-SENA (**■**), N-SENA (**♦**) and mix-SENA (★). The parameters of SENA detection are shown from the second top row to the bottom row as slope-5min, FC-5min, FC-10min and FCratio-10min/5min (abbreviated as *FCratio*), respectively and are all marked on their corresponding Y-axis. The RT-qPCR amplicons were all subject for NGS for verification (**Table S2**).

**Fig. S2.**
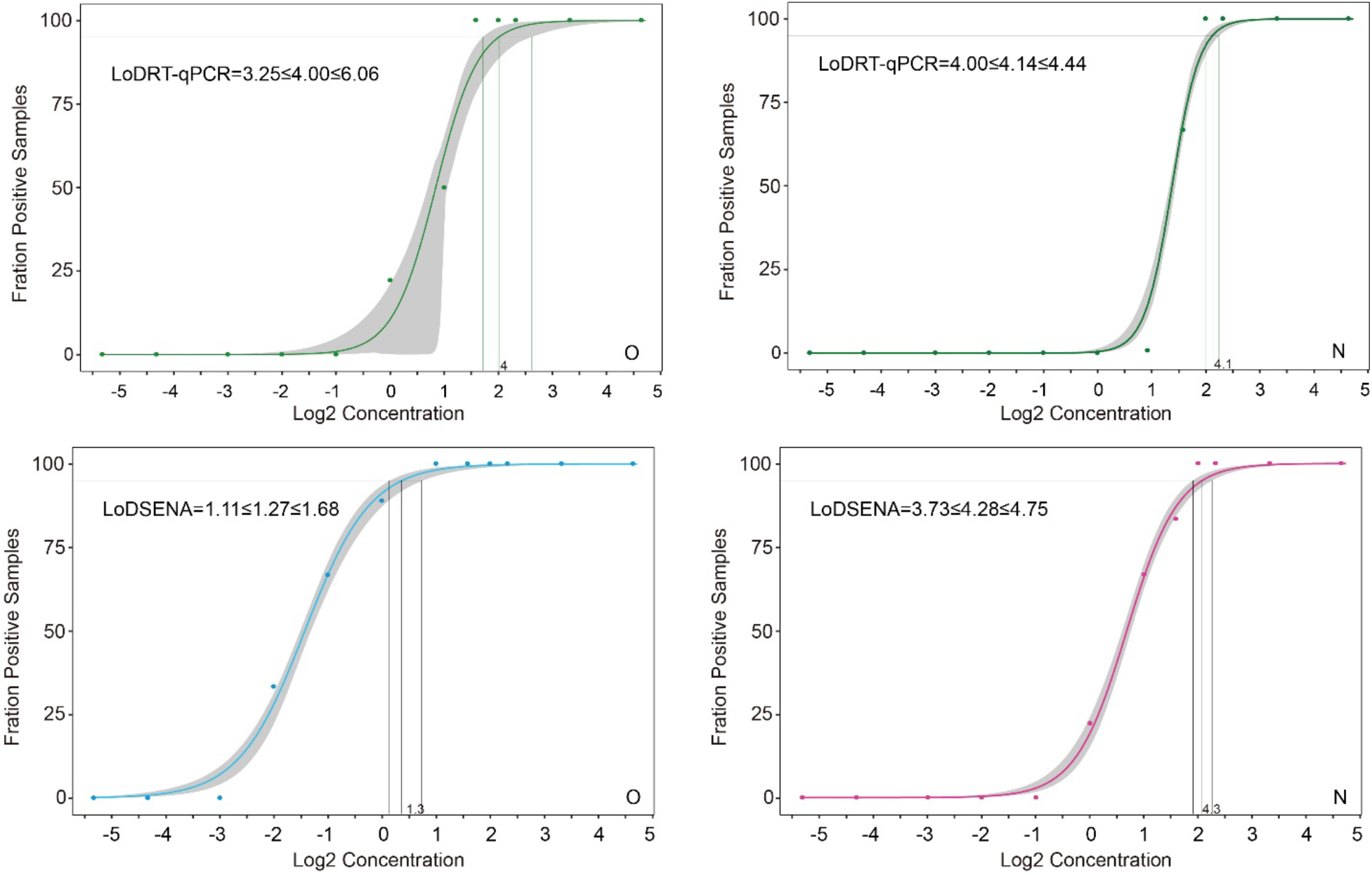
Determination of the LoD values with 95% CI for both RT-qPCR (O-Ct and N-Ct, both green dots) and SENA (*N-FCratio*, pink dots; and *O-FCratio*, blue dots) based on the systematic titration (Materials and Methods 4.4) All the experimental and analytical details are described in the text.

**Fig. S3.**
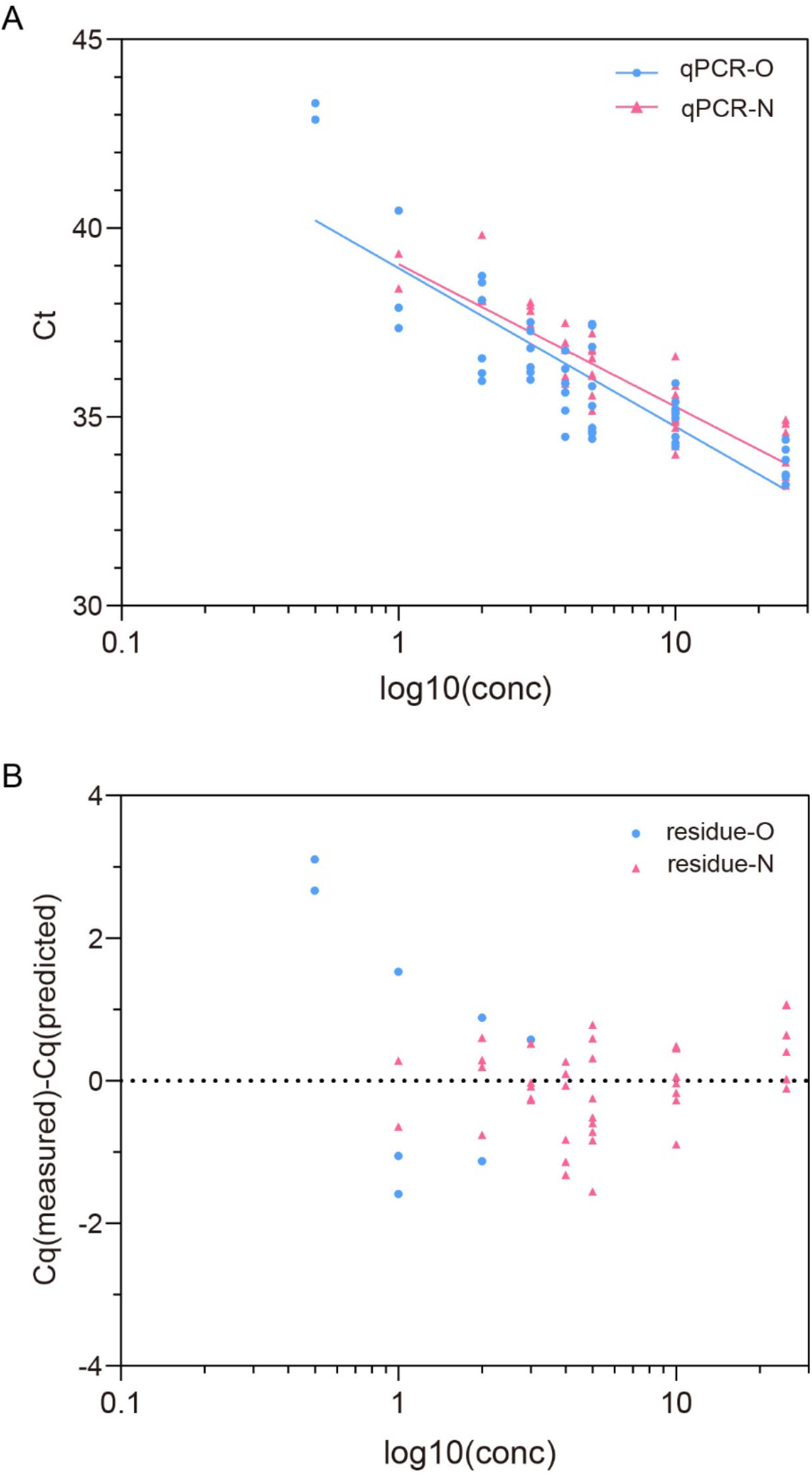
Linear regression (A) of the Ct values (● for ○ gene and ▲ for N gene) corresponding to the RT-qPCR titration experiment. The regression function for Ct (O) is: Y=38.94-4.198*log(x) (R^2^=0.7126) and the regression function for Ct (N) is: Y=39.05-3.776*log(x) (R^2^=0.7662), Panel B is the residue plot for these two regressions.

**Fig. S4.**
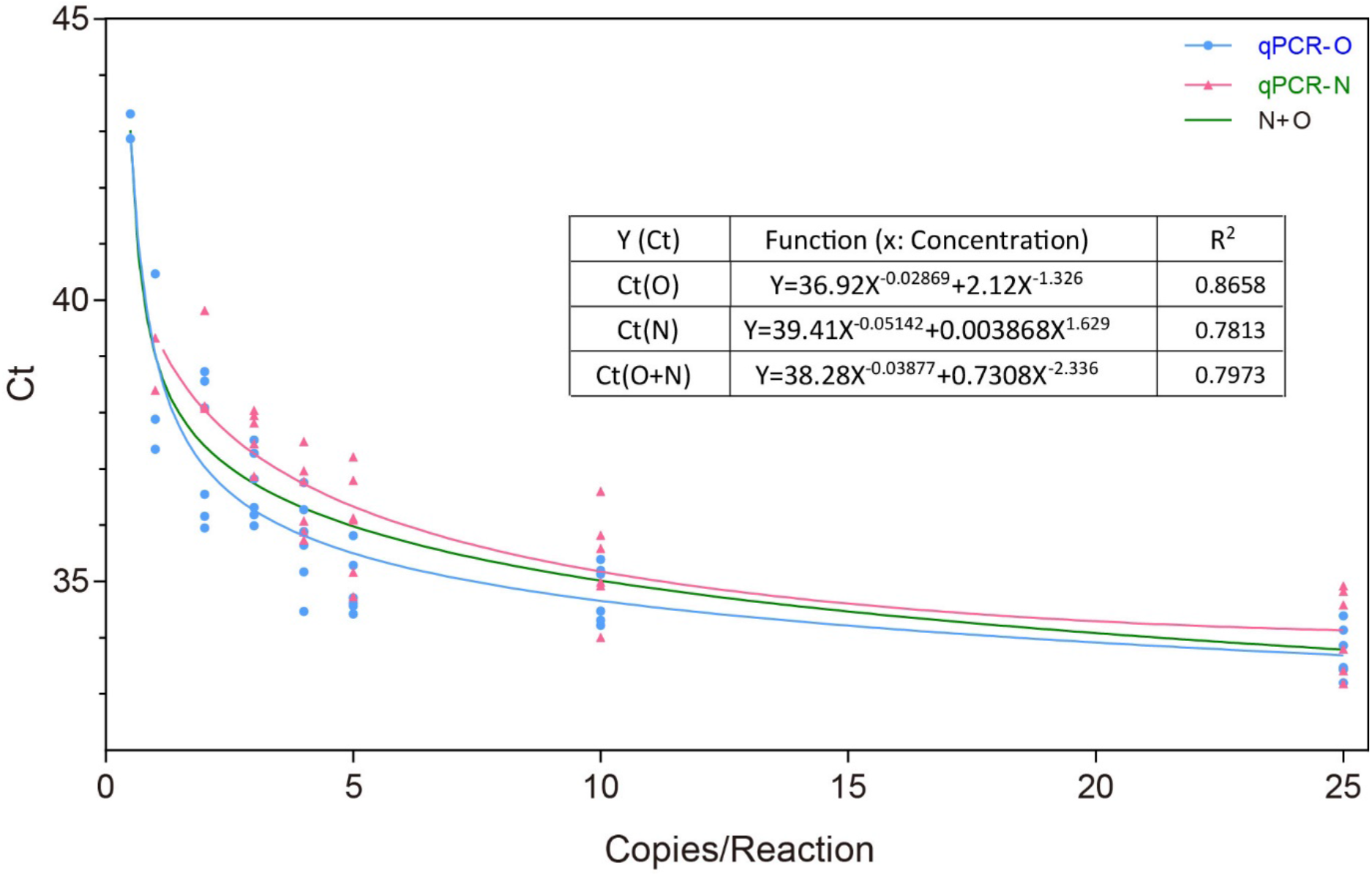
Regression of RT-qPCR Ct-values with the concentration of the templates (copies/Rx) The data is from **Table S2**. The regression functions for Ct-values of O gene (─), N gene (─) and both (─) (**Functions 1**) are illustrated as a table inside of the panel.

**Fig. S5.**
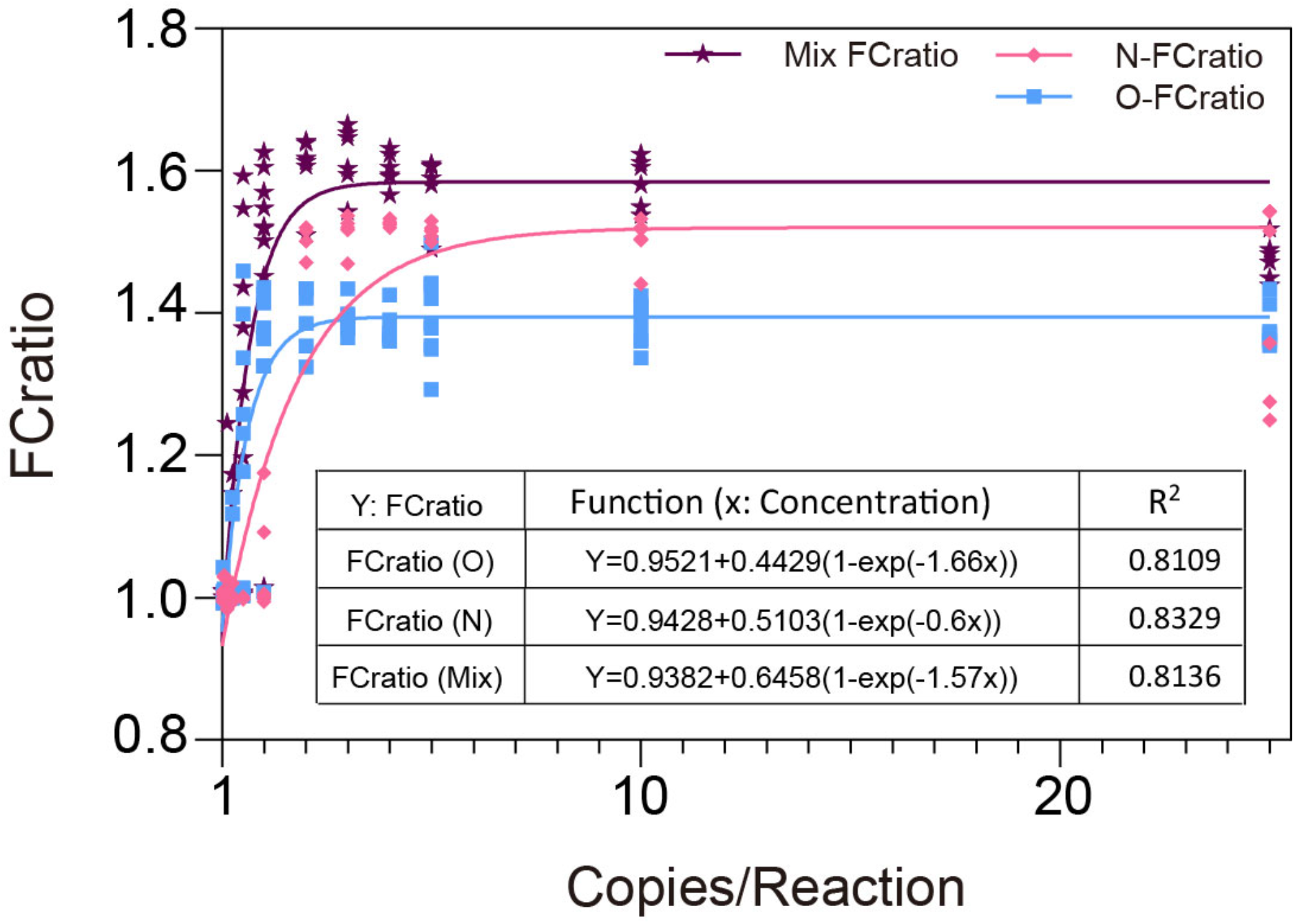
Regression of SENA *FCratio* with the concentration of the templates (copies/Rx) The data is from Table S2. The regression functions for *FCratios* of O-SENA (─), N-SENA (─) and mix-SENA (─) (**Functions 2**) are illustrated as a table inside of the panel.

**Table S1.**
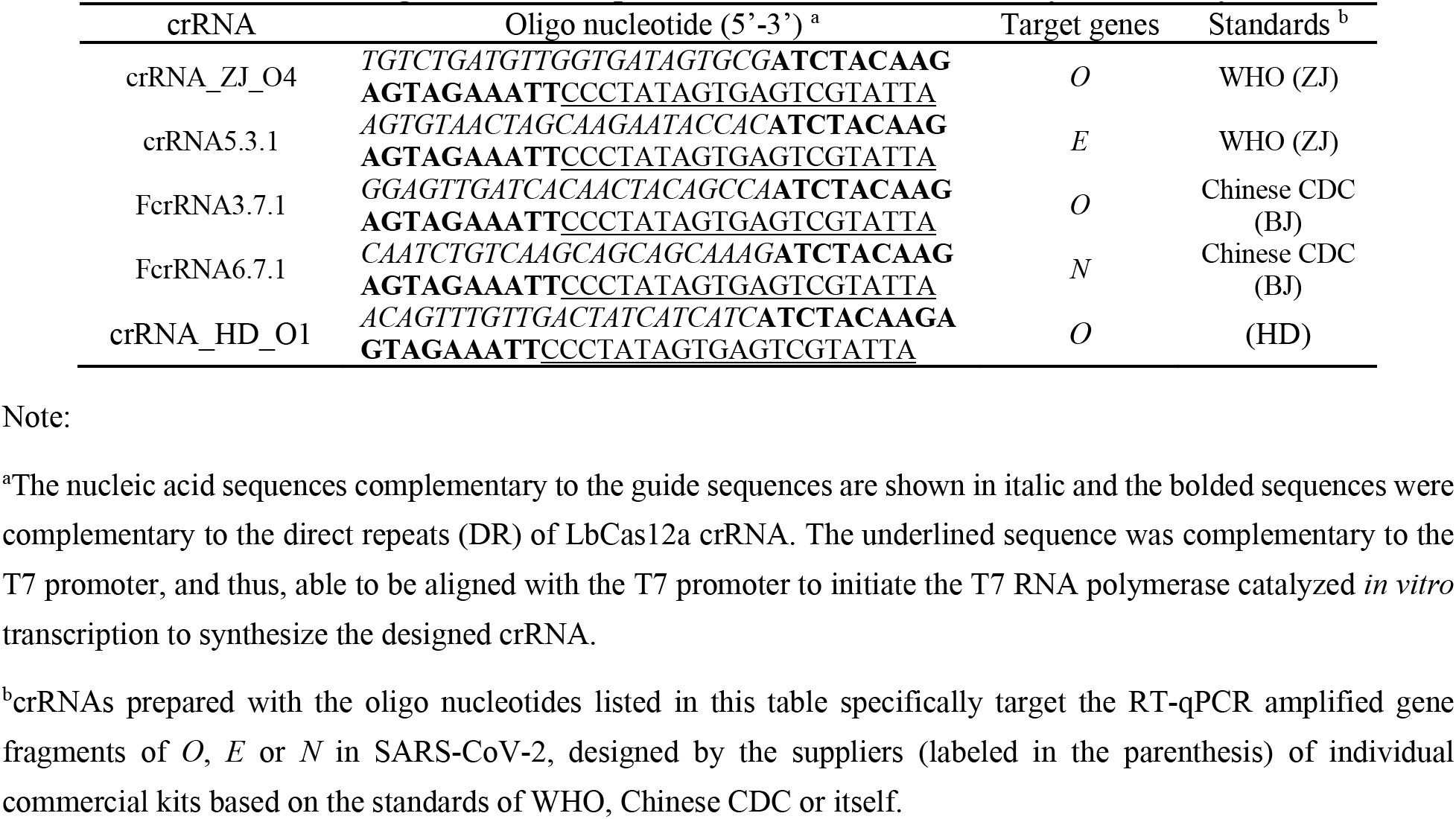
Oligo nucleotide sequences of crRNA for SENA assay in this study.

**Table S2-A.**
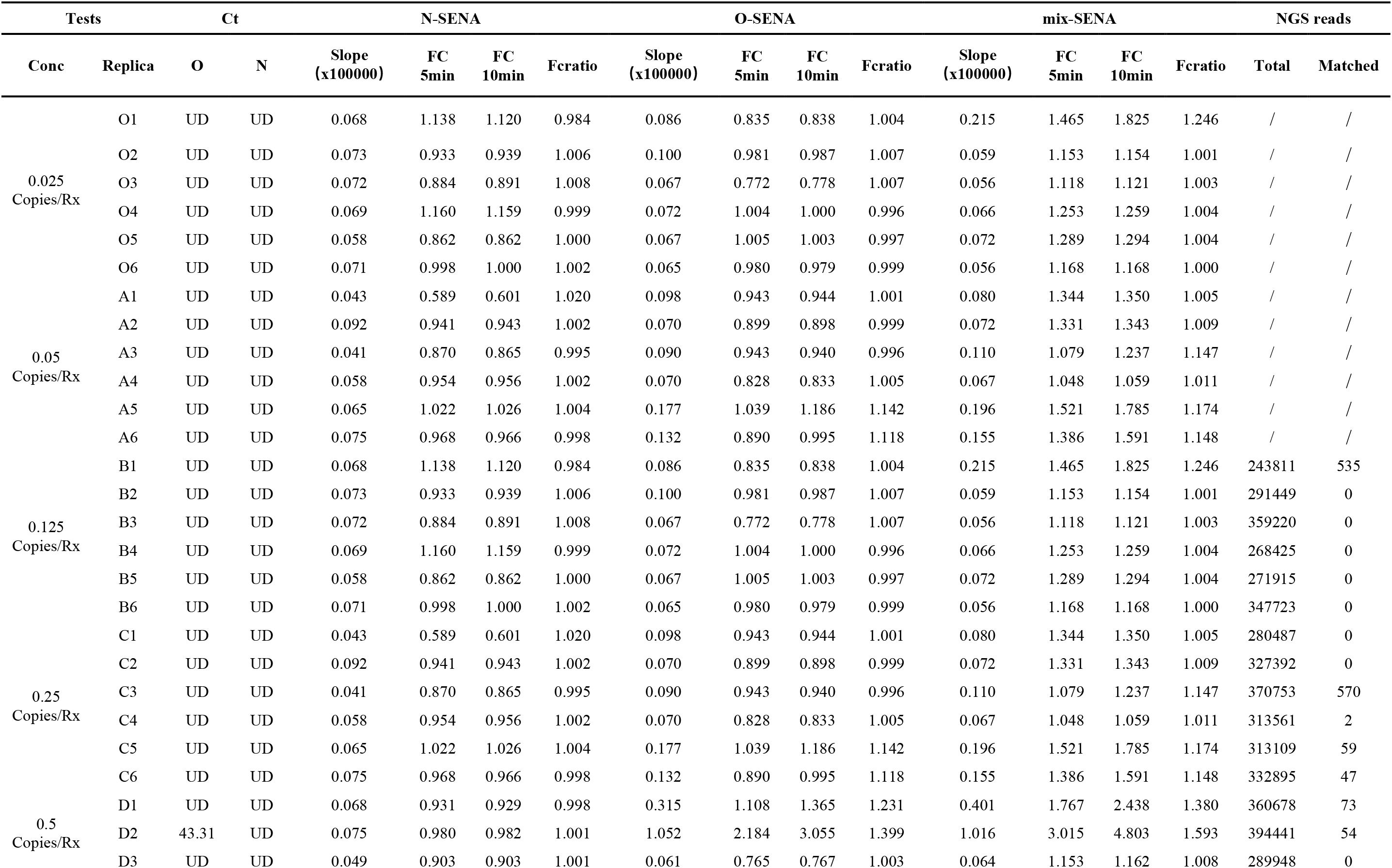
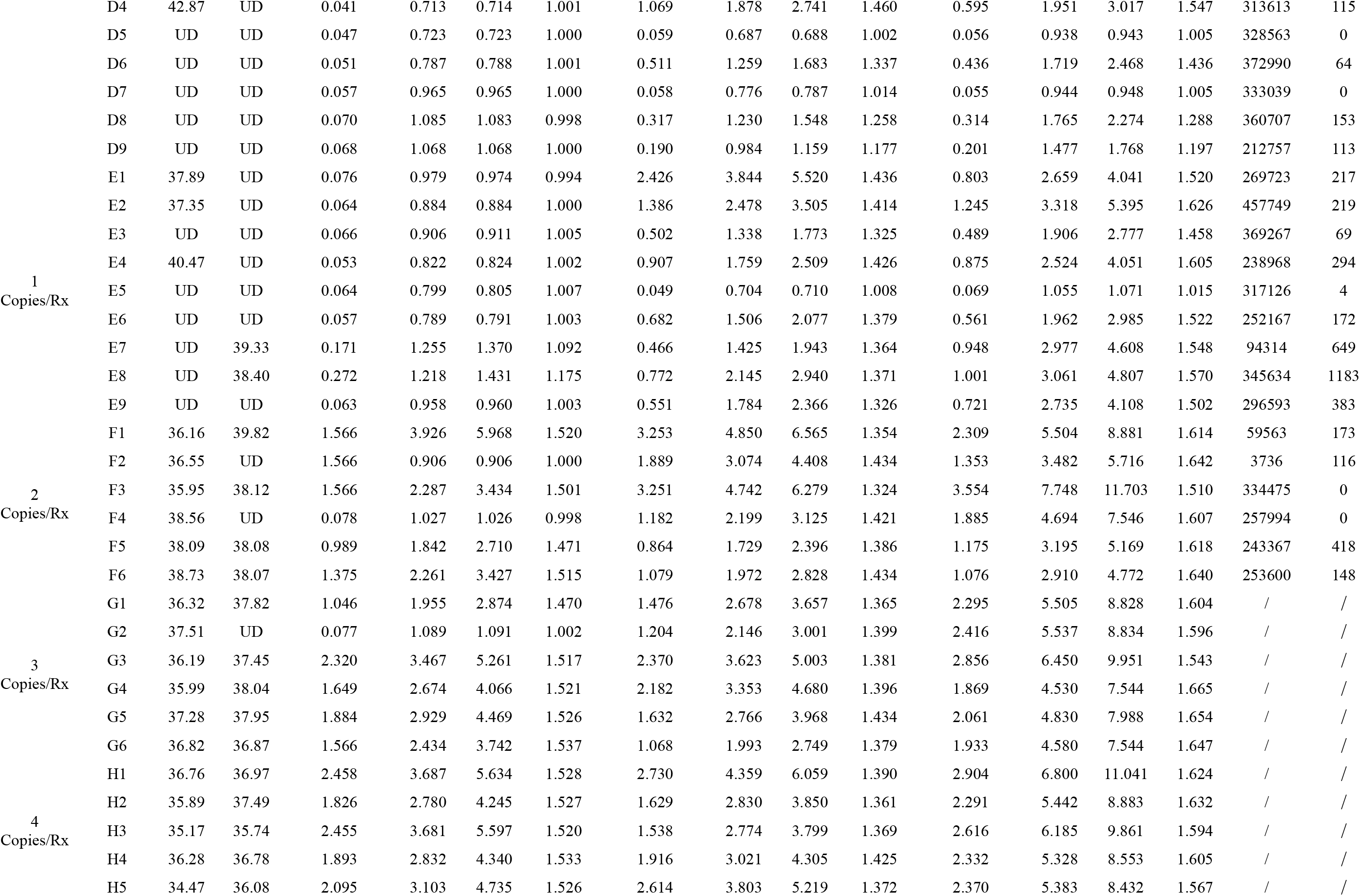
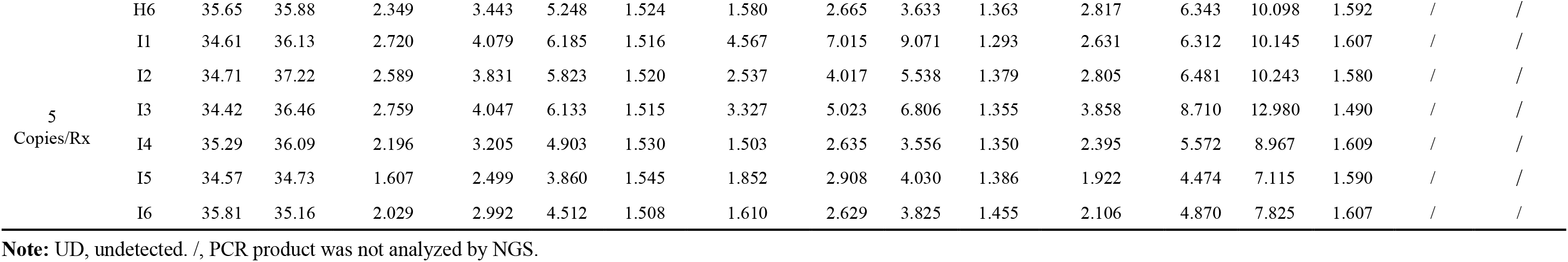
Original data of the titration experiment (Low template conc)

**Table S2-B.** Original data of the titration experiment (High template conc)

| Tests |  | Ct |  | N-SENA |  |  |  | O-SENA |  |  |  | mix-SENA |  |  |  | NGS reads |  |
| --- | --- | --- | --- | --- | --- | --- | --- | --- | --- | --- | --- | --- | --- | --- | --- | --- | --- |
| Conc | Replica | O | N | Slope<br>(x100000) | FC<br>5min | FC<br>10min | Fcratio | Slope<br>(x100000) | FC<br>5min | FC<br>10min | Fcratio | Slope<br>(x100000) | FC<br>5min | FC<br>10min | Fcratio | Total | Matched |
| 10<br>Copies/Rx | J1 | 34.23 | 34.01 | 0.528 | 3.700 | 5.625 | 1.520 | 3.394 | 5.168 | 6.910 | 1.337 | 2.232 | 5.499 | 8.931 | 1.624 | / | / |
|  | J2 | 34.31 | 36.61 | 0.528 | 3.898 | 5.857 | 1.503 | 2.679 | 4.355 | 5.923 | 1.360 | 2.921 | 6.863 | 10.553 | 1.538 | / | / |
|  | J3 | 34.47 | 34.99 | 0.528 | 3.163 | 4.802 | 1.518 | 2.412 | 3.913 | 5.495 | 1.404 | 2.146 | 5.230 | 8.432 | 1.612 | / | / |
|  | J4 | 35.39 | 35.59 | 2.785 | 4.042 | 6.083 | 1.505 | 2.817 | 4.434 | 6.047 | 1.364 | 2.966 | 6.814 | 10.559 | 1.550 | / | / |
|  | J5 | 35.14 | 35.82 | 2.130 | 3.131 | 4.801 | 1.533 | 1.794 | 3.069 | 4.321 | 1.408 | 2.258 | 5.290 | 8.360 | 1.580 | / | / |
|  | J6 | 35.45 | 34.92 | 2.637 | 1.273 | 1.526 | 1.442 | 1.539 | 3.772 | 5.327 | 1.424 | 1.834 | 4.311 | 6.923 | 1.606 | / | / |
| 25<br>Copies/Rx | H1 | 33.47 | 33.42 | 0.528 | 1.798 | 2.247 | 1.250 | 1.560 | 3.491 | 4.798 | 1.374 | 3.551 | 8.312 | 12.625 | 1.519 | / | / |
|  | H2 | 33.86 | 33.80 | 0.528 | 2.398 | 3.256 | 1.358 | 1.842 | 4.055 | 5.493 | 1.355 | 4.112 | 9.547 | 14.051 | 1.472 | / | / |
|  | H3 | 33.43 | 33.19 | 0.528 | 1.655 | 2.111 | 1.276 | 1.738 | 3.922 | 5.339 | 1.361 | 3.062 | 7.712 | 11.442 | 1.484 | / | / |
|  | H4 | 34.39 | 34.59 | 1.930 | 5.532 | 8.381 | 1.515 | 1.738 | 5.083 | 6.964 | 1.370 | 2.991 | 6.980 | 10.400 | 1.490 | / | / |
|  | H5 | 34.14 | 34.82 | 2.073 | 5.624 | 8.676 | 1.543 | 2.412 | 3.485 | 4.925 | 1.413 | 3.071 | 7.517 | 10.900 | 1.450 | / | / |
|  | H6 | 33.45 | 34.92 | 2.047 | 5.634 | 8.693 | 1.543 | 1.502 | 4.567 | 6.548 | 1.434 | 3.605 | 9.792 | 14.100 | 1.440 | / | / |

**Note:** /, PCR product was not analyzed by NGS.

**Table S3.** Comparison of SARS-CoV-2 measurement on clinical specimens by RT-qPCR and SENA

| Patient No. | Specimens | Clinical features |  | qPCR |  |  |  | SENA |  | NGS | Final outcome | Sample origin |
| --- | --- | --- | --- | --- | --- | --- | --- | --- | --- | --- | --- | --- |
|  |  | Symptoms | Diagnosis | Ct Value (E) | Ct Value (O) | Ct Value (N) | Interpretation | FCratio Value | Interpretation |  |  |  |
| P001 | Oropharyngeal swab | Fever, cough | COVID-19 suspected | N/A | 29.55 | 29.47 | + | 1.584 | + | / | Consistent Positive | SZII |
| P002 | Oropharyngeal swab | Fever, cough | COVID-19 suspected | N/A | 26.91 | 27.04 | + | 1.640 | + | / | Consistent Positive | SZII |
| P003 | Oropharyngeal swab | Fever, cough | COVID-19 suspected | N/A | 33.77 | 33.88 | + | 1.570 | + | / | Consistent Positive | SZII |
| P004 | Oropharyngeal swab | Fever, cough | COVID-19 suspected | N/A | 33.47 | 33.46 | + | 1.587 | + | / | Consistent Positive | SZII |
| P005 | Oropharyngeal swab | Fever, cough | COVID-19 suspected | N/A | 33.83 | 34.24 | + | 1.553 | + | / | Consistent Positive | SZII |
| P006 | Oropharyngeal swab | Fever, cough | COVID-19 suspected | N/A | 32.6 | 32.4 | + | 1.589 | + | / | Consistent Positive | SZII |
| P007 | Oropharyngeal swab | Fever, cough | COVID-19 suspected | N/A | 33.72 | 34.47 | + | 1.550 | + | / | Consistent Positive | SZII |
| P008 | Oropharyngeal swab | Fever, cough | COVID-19 suspected | N/A | 30.13 | 29.73 | + | 1.578 | + | / | Consistent Positive | SZII |
| P009 | Oropharyngeal swab | Fever, cough | COVID-19 suspected | N/A | 34.75 | 34.81 | + | 1.535 | + | / | Consistent Positive | SZII |
| P010 | Oropharyngeal swab | Fever, cough | COVID-19 suspected | N/A | 30.54 | 30.72 | + | 1.617 | + | / | Consistent Positive | SZII |
| P011 | Oropharyngeal swab | Asymptomatic | Close contacts | N/A | >40 | >40 | - | 1.003 | - | / | Consistent Negative | SZII |
| P012 | Oropharyngeal swab | Asymptomatic | Close contacts | N/A | >40 | >40 | - | 0.995 | - | / | Consistent Negative | SZII |
| P013 | Oropharyngeal swab | Asymptomatic | Close contacts | N/A | >40 | >40 | - | 0.998 | - | / | Consistent Negative | SZII |
| P014 | Oropharyngeal swab | Asymptomatic | Close contacts | N/A | >40 | >40 | - | 1.000 | - | / | Consistent Negative | SZII |
| P015 | Oropharyngeal swab | Asymptomatic | Close contacts | N/A | >40 | >40 | - | 0.996 | - | / | Consistent Negative | SZII |
| P016 | Oropharyngeal swab | Asymptomatic | Close contacts | N/A | >40 | >40 | - | 0.995 | - | / | Consistent Negative | SZII |
| P017 | Oropharyngeal swab | Asymptomatic | Close contacts | N/A | >40 | >40 | - | 0.993 | - | / | Consistent Negative | SZII |
| P018 | Oropharyngeal swab | Asymptomatic | Close contacts | N/A | >40 | >40 | - | 1.003 | - | / | Consistent Negative | SZII |
| P019 | Oropharyngeal swab | Asymptomatic | Close contacts | N/A | >40 | >40 | - | 0.995 | - | / | Consistent Negative | SZII |
| P020 | Oropharyngeal swab | Asymptomatic | Close contacts | N/A | >40 | >40 | - | 0.999 | - | / | Consistent Negative | SZII |
| P021 | Oropharyngeal swab | Asymptomatic | Close contacts | N/A | >40 | >40 | - | 0.995 | - | / | Consistent Negative | SZII |
| P022 | Oropharyngeal swab | Asymptomatic | Close contacts | N/A | >40 | >40 | - | 0.997 | - | / | Consistent Negative | SZII |
| P023 | Oropharyngeal swab | Asymptomatic | Close contacts | N/A | >40 | >40 | - | 0.994 | - | / | Consistent Negative | SZII |
| P024 | Oropharyngeal swab | Asymptomatic | Close contacts | N/A | >40 | >40 | - | 0.997 | - | / | Consistent Negative | SZII |
| P025 | Oropharyngeal swab | Asymptomatic | Close contacts | N/A | >40 | >40 | - | 0.996 | - | / | Consistent Negative | SZII |
| P026 | Oropharyngeal swab | Fever, cough | COVID-19 suspected | N/A | 18.00 | 20.00 | + | 1.447 | + | / | Consistent Positive | SZII |
| P027 | Oropharyngeal swab | Fever, cough | COVID-19 suspected | N/A | 26.00 | 27.50 | + | 1.290 | + | / | Consistent Positive | SZII |
| P028 | Oropharyngeal swab | Fever | COVID-19 suspected | N/A | >40 | 36.09 | + | 1.386 | + | + | Uncertain Positive | SZII |
| P029 | Oropharyngeal swab | Fever | COVID-19 suspected | N/A | >40 | 35.88 | + | 1.546 | + | + | Uncertain Positive | SZII |
| P030 | Oropharyngeal swab | Asymptomatic | Close contacts | N/A | >40 | >40 | - | 1.015 | - | - | Consistent Negative | SZII |
| P031 | Oropharyngeal swab | Asymptomatic | Close contacts | N/A | >40 | >40 | - | 1.017 | - | - | Consistent Negative | SZII |
| P032 | Oropharyngeal swab | Fever | COVID-19 suspected | N/A | >40 | 37.98 | + | 1.210 | + | + | Uncertain Positive | SZII |
| P033 | Oropharyngeal swab | Fever, Dyspnea | Pneumonia | N/A | >40 | >40 | - | 1.024 | - | - | Consistent Negative | SZII |
| P034 | Anal swabs | Fever, Dyspnea | Pneumonia | N/A | >40 | >40 | - | 1.012 | - | - | Consistent Negative | SZII |
| P035 | Oropharyngeal swab | Fever, Dyspnea | Pneumonia | N/A | >40 | >40 | - | 1.015 | - | - | Consistent Negative | SZII |
| P036 | Anal swabs | Fever, Dyspnea | Pneumonia | N/A | >40 | >40 | - | 1.008 | - | / | Consistent Negative | SZII |
| P037 | Oropharyngeal swab | Fever, Dyspnea | Pneumonia | N/A | >40 | >40 | - | 1.016 | - | - | Consistent Negative | SZII |
| P038 | Anal swabs | Fever, Dyspnea | Pneumonia | N/A | >40 | >40 | - | 1.022 | - | - | Consistent Negative | SZII |
| P039 | Oropharyngeal swab | Fever, Dyspnea | Pneumonia | N/A | >40 | >40 | - | 1.014 | - | - | Consistent Negative | SZII |
| P040 | Anal swabs | Fever, Dyspnea | Pneumonia | N/A | >40 | >40 | - | 1.014 | - | - | Consistent Negative | SZII |
| P041 | Oropharyngeal swab | Fever | COVID-19 suspected | N/A | >40 | >40 | - | 0.994 | - | / | Consistent Negative | SZII |
| P042 | Oropharyngeal swab | Fever | COVID-19 suspected | N/A | >40 | >40 | - | 1.011 | - | / | Consistent Negative | SZII |
| P043 | Oropharyngeal swab | Fever | COVID-19 suspected | N/A | >40 | >40 | - | 1.006 | - | / | Consistent Negative | SZII |
| P044 | Oropharyngeal swab | Fever | COVID-19 suspected | N/A | >40 | >40 | - | 1.018 | - | / | Consistent Negative | SZII |
| P045 | Oropharyngeal swab | Fever | COVID-19 suspected | N/A | >40 | >40 | - | 1.000 | - | / | Consistent Negative | SZII |
| P046 | Oropharyngeal swab | Fever | COVID-19 suspected | N/A | >40 | >40 | - | 0.988 | - | / | Consistent Negative | SZII |
| P047 | Oropharyngeal swab | Fever | COVID-19 suspected | N/A | >40 | >40 | - | 1.014 | - | / | Consistent Negative | SZII |
| P048 | Oropharyngeal swab | Fever | COVID-19 suspected | N/A | >40 | >40 | - | 1.002 | - | / | Consistent Negative | SZII |
| P049 | Oropharyngeal swab | Fever | COVID-19 suspected | N/A | >40 | >40 | - | 1.010 | - | / | Consistent Negative | SZII |
| P050 | Oropharyngeal swab | Fever | COVID-19 suspected | N/A | >40 | >40 | - | 1.006 | - | / | Consistent Negative | SZII |
| P051 | Oropharyngeal swab | Fever | COVID-19 suspected | N/A | >40 | >40 | - | 1.004 | - | / | Consistent Negative | SZII |
| P052 | Oropharyngeal swab | Fever | COVID-19 suspected | N/A | >40 | >40 | - | 0.998 | - | / | Consistent Negative | SZII |
| P053 | Oropharyngeal swab | Fever | COVID-19 suspected | N/A | >40 | >40 | - | 1.016 | - | / | Consistent Negative | SZII |
| P054 | Oropharyngeal swab | Fever | COVID-19 suspected | N/A | >40 | >40 | - | 0.998 | - | / | Consistent Negative | SZII |
| P055 | Oropharyngeal swab | Fever | COVID-19 suspected | N/A | 39.47 | >40 | Uncertain | 0.995 | - | - | Uncertain Negative | SZII |
| P056 | Oropharyngeal swab | Fever | COVID-19 suspected | N/A | >40 | >40 | - | 1.024 | - | / | Consistent Negative | SZII |
| P057 | Oropharyngeal swab | Fever | COVID-19 suspected | N/A | >40 | >40 | - | 1.008 | - | / | Consistent Negative | SZII |
| P058 | Oropharyngeal swab | Fever | COVID-19 suspected | N/A | >40 | >40 | - | 0.998 | - | / | Consistent Negative | SZII |
| P059 | Oropharyngeal swab | Fever | COVID-19 suspected | N/A | >40 | >40 | - | 0.995 | - | / | Consistent Negative | SZII |
| P060 | Oropharyngeal swab | Fever | COVID-19 suspected | N/A | >40 | >40 | - | 1.014 | - | / | Consistent Negative | SZII |
| P061 | Oropharyngeal swab | Fever | COVID-19 suspected | N/A | >40 | >40 | - | 1.010 | - | / | Consistent Negative | SZII |
| P062 | Oropharyngeal swab | Fever | COVID-19 suspected | N/A | >40 | >40 | - | 1.008 | - | / | Consistent Negative | SZII |
| P063 | Oropharyngeal swab | Fever | COVID-19 suspected | N/A | >40 | >40 | - | 1.007 | - | / | Consistent Negative | SZII |
| P064 | Oropharyngeal swab | Fever | COVID-19 suspected | N/A | >40 | >40 | - | 1.005 | - | / | Consistent Negative | SZII |
| P065 | Oropharyngeal swab | Fever | COVID-19 suspected | N/A | >40 | >40 | - | 1.016 | - | / | Consistent Negative | SZII |
| P066 | Oropharyngeal swab | Fever | COVID-19 suspected | N/A | >40 | >40 | - | 1.001 | - | / | Consistent Negative | SZII |
| P067 | Oropharyngeal swab | Fever | COVID-19 suspected | N/A | >40 | >40 | - | 1.001 | - | / | Consistent Negative | SZII |
| P068 | Oropharyngeal swab | Fever | COVID-19 suspected | N/A | >40 | >40 | - | 1.009 | - | / | Consistent Negative | SZII |
| P069 | Oropharyngeal swab | Fever | COVID-19 suspected | N/A | >40 | >40 | - | 1.005 | - | / | Consistent Negative | SZII |
| P070 | Oropharyngeal swab | Fever | COVID-19 suspected | N/A | >40 | >40 | - | 1.009 | - | / | Consistent Negative | SZII |
| P071 | Oropharyngeal swab | Fever | COVID-19 suspected | N/A | >40 | >40 | - | 1.003 | - | / | Consistent Negative | SZII |
| P072 | Oropharyngeal swab | Fever | COVID-19 suspected | N/A | >40 | >40 | - | 0.952 | - | / | Consistent Negative | SZII |
| P073 | Oropharyngeal swab | Fever | COVID-19 suspected | N/A | >40 | >40 | - | 1.009 | - | / | Consistent Negative | SZII |
| P074 | Oropharyngeal swab | Fever | COVID-19 suspected | N/A | >40 | >40 | - | 1.004 | - | / | Consistent Negative | SZII |
| P075 | Oropharyngeal swab | Fever | COVID-19 suspected | N/A | >40 | >40 | - | 0.992 | - | / | Consistent Negative | SZII |
| P076 | Oropharyngeal swab | Fever | COVID-19 suspected | N/A | >40 | >40 | - | 0.992 | - | / | Consistent Negative | SZII |
| P077 | Oropharyngeal swab | Fever | COVID-19 suspected | N/A | >40 | >40 | - | 1.000 | - | / | Consistent Negative | SZII |
| P078 | Oropharyngeal swab | Fever | COVID-19 suspected | N/A | >40 | >40 | - | 0.996 | - | / | Consistent Negative | SZII |
| P079 | Oropharyngeal swab | Fever | COVID-19 suspected | N/A | >40 | >40 | - | 0.995 | - | / | Consistent Negative | SZII |
| P080 | Oropharyngeal swab | Fever | COVID-19 suspected | N/A | >40 | >40 | - | 0.987 | - | / | Consistent Negative | SZII |
| P081 | Oropharyngeal swab | Fever | COVID-19 suspected | N/A | >40 | >40 | - | 1.009 | - | / | Consistent Negative | SZII |
| P082 | Oropharyngeal swab | Fever | COVID-19 suspected | N/A | >40 | >40 | - | 1.010 | - | / | Consistent Negative | SZII |
| P083 | Oropharyngeal swab | Fever | COVID-19 suspected | N/A | >40 | >40 | - | 0.999 | - | / | Consistent Negative | SZII |
| P084 | Oropharyngeal swab | Fever | COVID-19 suspected | N/A | >40 | >40 | - | 0.983 | - | / | Consistent Negative | SZII |
| P085 | Oropharyngeal swab | Fever | COVID-19 suspected | N/A | >40 | >40 | - | 1.011 | - | / | Consistent Negative | SZII |
| P086 | Oropharyngeal swab | Fever | COVID-19 suspected | N/A | >40 | >40 | - | 0.986 | - | / | Consistent Negative | SZII |
| P087 | Oropharyngeal swab | Fever | COVID-19 suspected | N/A | >40 | >40 | - | 0.990 | - | / | Consistent Negative | SZII |
| P088 | Oropharyngeal swab | Fever | COVID-19 suspected | N/A | >40 | >40 | - | 1.002 | - | / | Consistent Negative | SZII |
| P089 | Oropharyngeal swab | Fever | COVID-19 suspected | N/A | >40 | >40 | - | 1.002 | - | / | Consistent Negative | SZII |
| P090 | Oropharyngeal swab | Fever | COVID-19 suspected | N/A | >40 | >40 | - | 1.001 | - | / | Consistent Negative | SZII |
| P091 | Oropharyngeal swab | Fever | COVID-19 suspected | N/A | 39.70 | >40 | Uncertain | 0.992 | - | - | Uncertain Negative | SZII |
| P092 | Oropharyngeal swab | Fever | COVID-19 suspected | N/A | >40 | >40 | - | 1.001 | - | / | Consistent Negative | SZII |
| P093 | Oropharyngeal swab | Fever | COVID-19 suspected | N/A | >40 | >40 | - | 1.004 | - | / | Consistent Negative | SZII |
| P094 | Oropharyngeal swab | Fever | COVID-19 suspected | N/A | >40 | >40 | - | 0.999 | - | / | Consistent Negative | SZII |
| P095 | Oropharyngeal swab | Fever | COVID-19 suspected | N/A | >40 | >40 | - | 1.008 | - | / | Consistent Negative | SZII |
| P096 | Oropharyngeal swab | Fever | COVID-19 suspected | N/A | >40 | >40 | - | 1.006 | - | / | Consistent Negative | SZII |
| P097 | Oropharyngeal swab | Fever | COVID-19 suspected | N/A | >40 | >40 | - | 1.003 | - | / | Consistent Negative | SZII |
| P098 | Oropharyngeal swab | Fever | COVID-19 suspected | N/A | >40 | >40 | - | 1.010 | - | / | Consistent Negative | SZII |
| P099 | Oropharyngeal swab | Fever | COVID-19 suspected | N/A | >40 | >40 | - | 1.001 | - | / | Consistent Negative | SZII |
| P100 | Oropharyngeal swab | Fever | COVID-19 suspected | N/A | >40 | >40 | - | 1.006 | - | / | Consistent Negative | SZII |
| P101 | Oropharyngeal swab | Fever | COVID-19 suspected | N/A | >40 | >40 | - | 0.997 | - | / | Consistent Negative | SZII |
| P102 | Oropharyngeal swab | Fever | COVID-19 suspected | N/A | >40 | >40 | - | 1.004 | - | / | Consistent Negative | SZII |
| P103 | Oropharyngeal swab | Fever | COVID-19 suspected | N/A | 40.56 | >40 | Uncertain | 0.996 | - | - | Uncertain Negative | SZII |
| P104 | Oropharyngeal swab | Fever | COVID-19 suspected | N/A | >40 | >40 | - | 0.988 | - | / | Consistent Negative | SZII |
| P105 | Oropharyngeal swab | Fever | COVID-19 suspected | N/A | >40 | >40 | - | 1.001 | - | / | Consistent Negative | SZII |
| P106 | Oropharyngeal swab | Fever | COVID-19 suspected | N/A | >40 | >40 | - | 1.002 | - | / | Consistent Negative | SZII |
| P107 | Oropharyngeal swab | Fever | COVID-19 suspected | N/A | >40 | >40 | - | 1.000 | - | / | Consistent Negative | SZII |
| P108 | Oropharyngeal swab | Fever | COVID-19 suspected | N/A | >40 | >40 | - | 1.008 | - | / | Consistent Negative | SZII |
| P109 | Oropharyngeal swab | Fever | COVID-19 suspected | N/A | >40 | >40 | - | 1.005 | - | / | Consistent Negative | SZII |
| P110 | Oropharyngeal swab | Fever | COVID-19 suspected | N/A | >40 | >40 | - | 1.031 | - | / | Consistent Negative | SZII |
| P111 | Oropharyngeal swab | Fever | COVID-19 suspected | N/A | >40 | >40 | - | 1.004 | - | / | Consistent Negative | SZII |
| P112 | Oropharyngeal swab | Fever | COVID-19 suspected | N/A | >40 | >40 | - | 1.008 | - | / | Consistent Negative | SZII |
| P113 | Oropharyngeal swab | Fever | COVID-19 suspected | N/A | >40 | >40 | - | 1.013 | - | / | Consistent Negative | SZII |
| P114 | Oropharyngeal swab | Fever | COVID-19 suspected | N/A | >40 | >40 | - | 0.994 | - | / | Consistent Negative | SZII |
| P115 | Oropharyngeal swab | Fever | COVID-19 suspected | N/A | >40 | >40 | - | 0.998 | - | / | Consistent Negative | SZII |
| P116 | Oropharyngeal swab | Fever | COVID-19 suspected | N/A | >40 | >40 | - | 1.000 | - | / | Consistent Negative | SZII |
| P117 | Oropharyngeal swab | Fever | COVID-19 suspected | N/A | >40 | >40 | - | 1.000 | - | / | Consistent Negative | SZII |
| P118 | Oropharyngeal swab | Fever | COVID-19 suspected | N/A | >40 | >40 | - | 1.005 | - | / | Consistent Negative | SZII |
| P119 | Oropharyngeal swab | Fever | COVID-19 suspected | N/A | >40 | >40 | - | 0.992 | - | / | Consistent Negative | SZII |
| P120 | Oropharyngeal swab | Fever | COVID-19 suspected | N/A | >40 | >40 | - | 0.981 | - | / | Consistent Negative | SZII |
| P121 | Oropharyngeal swab | Fever | COVID-19 suspected | N/A | >40 | >40 | - | 1.015 | - | / | Consistent Negative | SZII |
| P122 | Oropharyngeal swab | Fever | COVID-19 suspected | N/A | >40 | >40 | - | 1.001 | - | / | Consistent Negative | SZII |
| P123 | Oropharyngeal swab | Fever | COVID-19 suspected | N/A | >40 | >40 | - | 1.001 | - | / | Consistent Negative | SZII |
| P124 | Oropharyngeal swab | Fever | COVID-19 suspected | N/A | >40 | >40 | - | 1.003 | - | / | Consistent Negative | SZII |
| P125 | Oropharyngeal swab | Fever | COVID-19 suspected | N/A | >40 | >40 | - | 1.000 | - | / | Consistent Negative | SZII |
| P126 | Oropharyngeal swab | Fever | COVID-19 suspected | N/A | >40 | >40 | - | 0.987 | - | / | Consistent Negative | SZII |
| P127 | Oropharyngeal swab | Fever | COVID-19 suspected | N/A | >40 | >40 | - | 0.994 | - | / | Consistent Negative | SZII |
| P128 | Oropharyngeal swab | Fever | COVID-19 suspected | N/A | >40 | >40 | - | 0.989 | - | / | Consistent Negative | SZII |
| P129 | Oropharyngeal swab | Fever | COVID-19 suspected | N/A | >40 | >40 | - | 0.978 | - | / | Consistent Negative | SZII |
| P130 | Oropharyngeal swab | Fever | COVID-19 suspected | N/A | >40 | >40 | - | 0.991 | - | / | Consistent Negative | SZII |
| P131 | Oropharyngeal swab | Fever | COVID-19 suspected | N/A | >40 | >40 | - | 1.001 | - | / | Consistent Negative | SZII |
| P132 | Oropharyngeal swab | Fever | COVID-19 suspected | N/A | >40 | >40 | - | 1.001 | - | / | Consistent Negative | SZII |
| P133 | Oropharyngeal swab | Fever | COVID-19 suspected | N/A | >40 | >40 | - | 0.998 | - | / | Consistent Negative | SZII |
| P134 | Oropharyngeal swab | Fever | COVID-19 suspected | N/A | >40 | >40 | - | 0.995 | - | / | Consistent Negative | SZII |
| P135 | Oropharyngeal swab | Fever, cough | COVID-19 suspected | N/A | 26.72 | 26.41 | + | 1.502 | + | / | Consistent Positive | SZII |
| P136 | Oropharyngeal swab | Fever, cough | COVID-19 suspected | N/A | 24.38 | 24.02 | + | 1.447 | + | / | Consistent Positive | SZII |
| P137 | Oropharyngeal swab | Fever, cough | COVID-19 suspected | N/A | 38.87 | 36.28 | Uncertain | 1.581 | + | / | Uncertain Positive | SZII |
| P138 | Oropharyngeal swab | Fever, cough | COVID-19 suspected | N/A | 39.22 | 38.21 | Uncertain | 1.609 | + | / | Uncertain Positive | SZII |
| P139 | Oropharyngeal swab | Fever, cough | COVID-19 suspected | N/A | 36.26 | 33.90 | + | 1.646 | + | / | Consistent Positive | SZII |
| P140-1 | Throat swab | No common respiratory symptoms | Came from Wuhan, suspected | N/A | >40 | >40 | - | 0.992 | - | / | Consistent Negative | DF |
| P140-2 | Nasopharyngeal swab (Left) | No common respiratory symptoms | Came from Wuhan, suspected | N/A | >40 | >40 | - | 1.003 | - | / | Consistent Negative | DF |
| P140-3 | Nasopharyngeal swab (Right) | No common respiratory symptoms | Came from Wuhan, suspected | N/A | 35.96 | 37.24 | + | 1.430 | + | + | Consistent Positive | DF |
| P140-4 | Blood serum | No common respiratory symptoms | Came from Wuhan, suspected | N/A | >40 | >40 | - | 1.002 | - | / | Consistent Negative | DF |
| P140-5 | Nasopharyngeal swab | No common respiratory symptoms | Came from Wuhan, suspected | N/A | >40 | >40 | - | 1.006 | - | / | Consistent Negative | DF |
| P140-6 | Nasol swab | No common respiratory symptoms | Came from Wuhan, suspected | N/A | >40 | >40 | - | 1.614 | + | + | False Negative | DF |
| P140-7 | Nasol swab | No common respiratory symptoms | Came from Wuhan, suspected | N/A | >40 | >40 | - | 1.369 | + | + | False Negative | DF |
| P140-8 | Blood serum | No common respiratory symptoms | Came from Wuhan, suspected | N/A | >40 | >40 | - | 1.000 | - | / | Consistent Negative | DF |
| P140-9 | Nasopharyngeal swab (Left) | No common respiratory symptoms | Came from Wuhan, suspected | N/A | >40 | >40 | - | 1.004 | - | - | Consistent Negative | DF |
| P140-10 | Nasopharyngeal swab (Right) | No common respiratory symptoms | Came from Wuhan, suspected | N/A | >40 | >40 | - | 1.004 | - | / | Consistent Negative | DF |
| P140-11 | Throat swab | No common respiratory symptoms | Came from Wuhan, suspected | N/A | >40 | >40 | - | 1.002 | - | / | Consistent Negative | DF |
| P140-12 | Serum | No common respiratory symptoms | Came from Wuhan, suspected | N/A | >40 | >40 | - | 1.007 | - | / | Consistent Negative | DF |
| P140-13 | Plasma | No common respiratory symptoms | Came from Wuhan, suspected | N/A | >40 | >40 | - | 1.005 | - | / | Consistent Negative | DF |
| P140-14 | Fecal | No common respiratory symptoms | Came from Wuhan, suspected | N/A | >40 | >40 | - | 1.002 | - | / | Consistent Negative | DF |
| P141 | Nasopharyngeal swab | Fever | COVID-19 suspected | 32.68 | 33.98 | 33.42 | + | 1.673 | + | + | Consistent Positive | RJ |
| P142 | Nasopharyngeal swab | Fever | COVID-19 suspected | 35.76 | 37.54 | 36.28 | + | 1.391 | + | + | Consistent Positive | RJ |
| P143 | Nasopharyngeal swab | Fever, Chest pain | COVID-19 suspected | >45 | >45 | >45 | - | 1.245 | + | + | False Negative | RJ |
| P144 | Nasopharyngeal swab | Cough | Upper respiratory infection | 41.92 | >45 | >45 | Uncertain | 1.022 | - | - | Uncertain Negative | RJ |
| P145 | Nasopharyngeal swab | Fever | Upper respiratory infection | 41.42 | >45 | >45 | Uncertain | 0.897 | - | - | Uncertain Negative | RJ |
| P146 | Nasopharyngeal swab | Chest pain | COVID-19 suspected | 42.62 | >45 | >45 | Uncertain | 0.898 | - | - | Uncertain Negative | RJ |
| P147 | Nasopharyngeal swab | Cough | Upper respiratory infection | 41.34 | >45 | >45 | Uncertain | 0.730 | - | - | Uncertain Negative | RJ |
| P148 | Nasopharyngeal swab | Fever | COVID-19 suspected | 42.89 | >45 | >45 | Uncertain | 0.812 | - | - | Uncertain Negative | RJ |
| P149 | Nasopharyngeal swab | Dyspnea | Pneumonia | 42.65 | >45 | >45 | Uncertain | 0.768 | - | - | Uncertain Negative | RJ |
| P150 | Nasopharyngeal swab | Fever | COVID-19 suspected | 41.82 | >45 | >45 | Uncertain | 0.991 | - | - | Uncertain Negative | RJ |
| P151 | Nasopharyngeal swab | Asymptomatic | Upper respiratory infection | 42.14 | >45 | >45 | Uncertain | 0.960 | - | - | Uncertain Negative | RJ |
| P152 | Nasopharyngeal swab | Dyspnea | COVID-19 suspected | 41.74 | >45 | >45 | Uncertain | 0.960 | - | - | Uncertain Negative | RJ |
| P153 | Nasopharyngeal swab | Fever | Upper respiratory infection | 41.84 | >45 | >45 | Uncertain | 0.897 | - | - | Uncertain Negative | RJ |
| P154 | Nasopharyngeal swab | Chest pain | Heart failure | 42.04 | >45 | >45 | Uncertain | 0.991 | - | - | Uncertain Negative | RJ |
| P155 | Nasopharyngeal swab | cough | Pneumonia | 41.12 | >45 | >45 | Uncertain | 0.898 | - | - | Uncertain Negative | RJ |
| P156 | Nasopharyngeal swab | Fever | Pneumonia | >45 | >45 | >45 | - | 0.936 | - | / | Consistent Negative | RJ |
| P157 | Nasopharyngeal swab | Cough | Upper respiratory infection | >45 | >45 | >45 | - | 0.894 | - | / | Consistent Negative | RJ |
| P158 | Nasopharyngeal swab | Fever | COVID-19 suspected | >45 | >45 | >45 | - | 0.970 | - | / | Consistent Negative | RJ |
| P159 | Nasopharyngeal swab | Fever | COVID-19 suspected | 44.62 | >45 | >45 | - | 0.870 | - | / | Consistent Negative | RJ |
| P160 | Nasopharyngeal swab | Dyspnea | Diabetes | >45 | >45 | >45 | - | 0.817 | - | / | Consistent Negative | RJ |
| P161 | Nasopharyngeal swab | Cough | Upper respiratory infection | >45 | >45 | >45 | - | 0.804 | - | / | Consistent Negative | RJ |
| P162 | Nasopharyngeal swab | Fever | Diabetes | >45 | >45 | >45 | - | 0.859 | - | / | Consistent Negative | RJ |
| P163 | Nasopharyngeal swab | Dyspnea | Upper respiratory infection | >45 | >45 | >45 | - | 0.815 | - | / | Consistent Negative | RJ |
| P164 | Nasopharyngeal swab | Fever | Upper respiratory infection | >45 | >45 | >45 | - | 0.871 | - | / | Consistent Negative | RJ |
| P165 | Nasopharyngeal swab | Cough | Diabetes | >45 | >45 | >45 | - | 0.802 | - | / | Consistent Negative | RJ |
| P166 | Nasopharyngeal swab | Cough | Diabetes | >45 | >45 | >45 | - | 0.990 | - | / | Consistent Negative | RJ |
| P167 | Nasopharyngeal swab | Fever | Upper respiratory infection | >45 | >45 | >45 | - | 0.817 | - | / | Consistent Negative | RJ |
| P168 | Nasopharyngeal swab | Fever | Diabetes | >45 | >45 | >45 | - | 0.760 | - | / | Consistent Negative | RJ |
| P169 | Nasopharyngeal swab | Dyspnea | Upper respiratory infection | >45 | >45 | >45 | - | 0.802 | - | / | Consistent Negative | RJ |
| P170 | Nasopharyngeal swab | Asymptomatic | Close contacts | >45 | >45 | >45 | - | 0.636 | - | / | Consistent Negative | RJ |
| P171 | Nasopharyngeal swab | Fever | Upper respiratory infection | >45 | >45 | >45 | - | 0.758 | - | / | Consistent Negative | RJ |
| P172 | Nasopharyngeal swab | Fever | Upper respiratory infection | 43.12 | >45 | >45 | - | 0.931 | - | / | Consistent Negative | RJ |
| P173 | Nasopharyngeal swab | Chest pain | Pneumonia | >45 | >45 | >45 | - | 0.758 | - | / | Consistent Negative | RJ |
| P174 | Nasopharyngeal swab | Fever | Diabetes | >45 | >45 | >45 | - | 0.951 | - | / | Consistent Negative | RJ |
| P175 | Nasopharyngeal swab | Cough | Upper respiratory infection | >45 | >45 | >45 | - | 0.974 | - | / | Consistent Negative | RJ |
| P176 | Nasopharyngeal swab | Chest pain | Pneumonia | >45 | >45 | >45 | - | 0.683 | - | / | Consistent Negative | RJ |
| P177 | Nasopharyngeal swab | Dyspnea | Upper respiratory infection | 43.65 | >45 | >45 | - | 0.959 | - | / | Consistent Negative | RJ |
| P178 | Nasopharyngeal swab | Cough | Upper respiratory infection | >45 | >45 | >45 | - | 0.926 | - | / | Consistent Negative | RJ |
| P179 | Nasopharyngeal swab | Fever | COVID-19 suspected | 44.12 | >45 | >45 | - | 0.761 | - | / | Consistent Negative | RJ |
| P180 | Nasopharyngeal swab | Fever | COVID-19 suspected | >45 | >45 | >45 | - | 0.685 | - | / | Consistent Negative | RJ |
| P181 | Nasopharyngeal swab | Fever | Upper respiratory infection | >45 | >45 | >45 | - | 0.815 | - | / | Consistent Negative | RJ |
| P182 | Nasopharyngeal swab | Asymptomatic | Close contacts | >45 | >45 | >45 | - | 0.943 | - | / | Consistent Negative | RJ |
| P183 | Nasopharyngeal swab | Dyspnea | Pneumonia | >45 | >45 | >45 | - | 0.850 | - | / | Consistent Negative | RJ |
| P184 | Nasopharyngeal swab | Cough | Upper respiratory infection | >45 | >45 | >45 | - | 0.758 | - | / | Consistent Negative | RJ |
| P185 | Nasopharyngeal swab | Fever | Upper respiratory infection | >45 | >45 | >45 | - | 0.582 | - | / | Consistent Negative | RJ |
| P186 | Nasopharyngeal swab | Cough | Upper respiratory infection | >45 | >45 | >45 | - | 0.655 | - | / | Consistent Negative | RJ |
| P187 | Nasopharyngeal swab | Cough | Upper respiratory infection | >45 | >45 | >45 | - | 0.936 | - | / | Consistent Negative | RJ |
| P188 | Nasopharyngeal swab | Fever | Pneumonia | >45 | >45 | >45 | - | 0.647 | - | / | Consistent Negative | RJ |
| P189 | Nasopharyngeal swab | Fever | Upper respiratory infection | >45 | >45 | >45 | - | 0.699 | - | / | Consistent Negative | RJ |
| P190 | Nasopharyngeal swab | Fever | Pneumonia | >45 | >45 | >45 | - | 0.994 | - | / | Consistent Negative | RJ |
| P191 | Nasopharyngeal swab | Fever | Upper respiratory infection | >45 | >45 | >45 | - | 0.764 | - | / | Consistent Negative | RJ |
| P192 | Nasopharyngeal swab | Fever | Upper respiratory infection | >45 | >45 | >45 | - | 0.626 | - | / | Consistent Negative | RJ |
| P193 | Nasopharyngeal swab | Cough | Diabetes | >45 | >45 | >45 | - | 0.800 | - | / | Consistent Negative | RJ |
| P194 | Nasopharyngeal swab | Fever | Upper respiratory infection | >45 | >45 | >45 | - | 0.674 | - | / | Consistent Negative | RJ |
| P195 | Nasopharyngeal swab | Dyspnea | Pneumonia | >45 | >45 | >45 | - | 0.589 | - | / | Consistent Negative | RJ |
| P196 | Nasopharyngeal swab | Fever | Upper respiratory infection | >45 | >45 | >45 | - | 0.580 | - | / | Consistent Negative | RJ |
| P197 | Nasopharyngeal swab | Fever | Upper respiratory infection | >45 | >45 | >45 | - | 0.630 | - | / | Consistent Negative | RJ |
| P198 | Nasopharyngeal swab | Asymptomatic | Close contacts | >45 | >45 | >45 | - | 0.846 | - | / | Consistent Negative | RJ |
| P199 | Nasopharyngeal swab | Fever | Upper respiratory infection | >45 | >45 | >45 | - | 0.644 | - | / | Consistent Negative | RJ |
| P200 | Nasopharyngeal swab | Fever | Upper respiratory infection | >45 | >45 | >45 | - | 0.760 | - | / | Consistent Negative | RJ |
| P201 | Nasopharyngeal swab | Cough | Pneumonia | >45 | >45 | >45 | - | 0.949 | - | / | Consistent Negative | RJ |
| P202 | Nasopharyngeal swab | Fever | Upper respiratory infection | >45 | >45 | >45 | - | 0.905 | - | / | Consistent Negative | RJ |
| P203 | Nasopharyngeal swab | Fever | Pneumonia | >45 | >45 | >45 | - | 0.799 | - | / | Consistent Negative | RJ |
| P204 | Nasopharyngeal swab | Fever | Pneumonia | >45 | >45 | >45 | - | 0.905 | - | / | Consistent Negative | RJ |
| P205 | Nasopharyngeal swab | Asymptomatic | Close contacts | >45 | >45 | >45 | - | 0.868 | - | / | Consistent Negative | RJ |
| P206 | Nasopharyngeal swab | Fever | Upper respiratory infection | >45 | >45 | >45 | - | 0.674 | - | / | Consistent Negative | RJ |
| P207 | Nasopharyngeal swab | Asymptomatic | Close contacts | >45 | >45 | >45 | - | 0.936 | - | / | Consistent Negative | RJ |
| P208 | Nasopharyngeal swab | Chest pain | Pneumonia | >45 | >45 | >45 | - | 0.802 | - | / | Consistent Negative | RJ |
| P209 | Nasopharyngeal swab | Fever | Upper respiratory infection | >45 | >45 | >45 | - | 0.630 | - | / | Consistent Negative | RJ |
| P210 | Nasopharyngeal swab | Fever | COVID-19 suspected | >45 | >45 | >45 | - | 0.737 | - | / | Consistent Negative | RJ |
| P211 | Nasopharyngeal swab | Fever | COVID-19 suspected | >45 | >45 | >45 | - | 0.905 | - | / | Consistent Negative | RJ |
| P212 | Nasopharyngeal swab | Fever | Upper respiratory infection | >45 | >45 | >45 | - | 0.999 | - | / | Consistent Negative | RJ |
| P213 | Nasopharyngeal swab | Fever | Upper respiratory infection | >45 | >45 | >45 | - | 0.942 | - | / | Consistent Negative | RJ |
| P214 | Nasopharyngeal swab | Cough | Diabetes | >45 | >45 | >45 | - | 0.848 | - | / | Consistent Negative | RJ |
| P215 | Nasopharyngeal swab | Cough | Upper respiratory infection | >45 | >45 | >45 | - | 0.772 | - | / | Consistent Negative | RJ |
| P216 | Nasopharyngeal swab | Cough | Upper respiratory infection | >45 | >45 | >45 | - | 0.948 | - | / | Consistent Negative | RJ |
| P217 | Nasopharyngeal swab | Fever | Upper respiratory infection | >45 | >45 | >45 | - | 0.848 | - | / | Consistent Negative | RJ |
| P218 | Nasopharyngeal swab | Fever | Diabetes | >45 | >45 | >45 | - | 0.816 | - | / | Consistent Negative | RJ |
| P219 | Nasopharyngeal swab | Fever | Upper respiratory infection | >45 | >45 | >45 | - | 0.759 | - | / | Consistent Negative | RJ |
| P220 | Nasopharyngeal swab | Fever | COVID-19 suspected | >45 | >45 | >45 | - | 0.825 | - | / | Consistent Negative | RJ |
| P221 | Nasopharyngeal swab | Chest pain | Heart failure | >45 | >45 | >45 | - | 0.807 | - | / | Consistent Negative | RJ |
| P222 | Nasopharyngeal swab | Fever | Upper respiratory infection | >45 | >45 | >45 | - | 0.654 | - | / | Consistent Negative | RJ |
| P223 | Nasopharyngeal swab | Asymptomatic | Close contacts | >45 | >45 | >45 | - | 0.931 | - | / | Consistent Negative | RJ |
| P224 | Nasopharyngeal swab | Fever | Upper respiratory infection | >45 | >45 | >45 | - | 0.973 | - | / | Consistent Negative | RJ |
| P225 | Nasopharyngeal swab | Fever | Upper respiratory infection | >45 | >45 | >45 | - | 0.985 | - | / | Consistent Negative | RJ |
| P226 | Nasopharyngeal swab | Fever | Upper respiratory infection | >45 | >45 | >45 | - | 1.027 | - | / | Consistent Negative | RJ |
| P227 | Nasopharyngeal swab | Asymptomatic | Close contacts | >45 | >45 | >45 | - | 1.011 | - | / | Consistent Negative | RJ |
| P228 | Nasopharyngeal swab | Fever | Upper respiratory infection | 44.18 | >45 | >45 | - | 1.049 | - | / | Consistent Negative | RJ |
| P229 | Nasopharyngeal swab | Fever | Diabetes | >45 | >45 | >45 | - | 0.970 | - | / | Consistent Negative | RJ |
| P230 | Nasopharyngeal swab | Fever | Upper respiratory infection | >45 | >45 | >45 | - | 0.921 | - | / | Consistent Negative | RJ |
| P231 | Nasopharyngeal swab | Fever | COVID-19 suspected | >45 | >45 | >45 | - | 0.906 | - | / | Consistent Negative | RJ |
| P232 | Nasopharyngeal swab | Fever | Upper respiratory infection | >45 | >45 | >45 | - | 1.082 | - | / | Consistent Negative | RJ |
| P233 | Nasopharyngeal swab | Dyspnea | Pneumonia | >45 | >45 | >45 | - | 1.027 | - | / | Consistent Negative | RJ |
| P234 | Nasopharyngeal swab | Cough | Upper respiratory infection | >45 | >45 | >45 | - | 0.926 | - | / | Consistent Negative | RJ |
| P235 | Nasopharyngeal swab | Fever | Upper respiratory infection | >45 | >45 | >45 | - | 0.985 | - | / | Consistent Negative | RJ |
| P236 | Nasopharyngeal swab | Fever | COVID-19 suspected | >45 | >45 | >45 | - | 0.985 | - | / | Consistent Negative | RJ |
| P237 | Nasopharyngeal swab | Chest pain | Heart failure | >45 | >45 | >45 | - | 1.049 | - | / | Consistent Negative | RJ |
| P238 | Nasopharyngeal swab | Fever | Upper respiratory infection | >45 | >45 | >45 | - | 1.032 | - | / | Consistent Negative | RJ |
| P239 | Nasopharyngeal swab | Asymptomatic | Close contacts | >45 | >45 | >45 | - | 1.082 | - | / | Consistent Negative | RJ |
| P240 | Nasopharyngeal swab | Asymptomatic | Close contacts | >45 | >45 | >45 | - | 0.989 | - | / | Consistent Negative | RJ |
| P241 | Nasopharyngeal swab | Fever | Upper respiratory infection | >45 | >45 | >45 | - | 1.049 | - | / | Consistent Negative | RJ |
| P242 | Nasopharyngeal swab | Cough | Upper respiratory infection | >45 | >45 | >45 | - | 1.049 | - | / | Consistent Negative | RJ |
| P243 | Nasopharyngeal swab | Asymptomatic | Close contacts | >45 | >45 | >45 | - | 0.906 | - | / | Consistent Negative | RJ |
| P244 | Nasopharyngeal swab | Cough | Upper respiratory infection | >45 | >45 | >45 | - | 0.921 | - | / | Consistent Negative | RJ |
| P245 | Nasopharyngeal swab | Fever | Upper respiratory infection | >45 | >45 | >45 | - | 0.843 | - | / | Consistent Negative | RJ |
| P246 | Nasopharyngeal swab | Fever | Upper respiratory infection | >45 | >45 | >45 | - | 0.936 | - | / | Consistent Negative | RJ |
| P247 | Nasopharyngeal swab | Fever | Diabetes | >45 | >45 | >45 | - | 1.035 | - | / | Consistent Negative | RJ |
| P248 | Nasopharyngeal swab | Asymptomatic | Close contacts | >45 | >45 | >45 | - | 0.936 | - | / | Consistent Negative | RJ |
| P249 | Nasopharyngeal swab | Asymptomatic | Close contacts | >45 | >45 | >45 | - | 0.906 | - | / | Consistent Negative | RJ |
| P250 | Nasopharyngeal swab | Fever | COVID-19 suspected | >45 | >45 | >45 | - | 1.066 | - | / | Consistent Negative | RJ |
| P251 | Nasopharyngeal swab | Asymptomatic | Close contacts | >45 | >45 | >45 | - | 0.950 | - | / | Consistent Negative | RJ |
| P252 | Nasopharyngeal swab | Dyspnea | Upper respiratory infection | >45 | >45 | >45 | - | 0.970 | - | / | Consistent Negative | RJ |
| P253 | Nasopharyngeal swab | Asymptomatic | Close contacts | >45 | >45 | >45 | - | 0.921 | - | / | Consistent Negative | RJ |
| P254 | Nasopharyngeal swab | Cough | Upper respiratory infection | >45 | >45 | >45 | - | 1.027 | - | / | Consistent Negative | RJ |
| P255 | Nasopharyngeal swab | Fever | Upper respiratory infection | >45 | >45 | >45 | - | 1.032 | - | / | Consistent Negative | RJ |
| P256 | Nasopharyngeal swab | Fever | COVID-19 suspected | >45 | >45 | >45 | - | 0.936 | - | / | Consistent Negative | RJ |
| P257 | Nasopharyngeal swab | Asymptomatic | Close contacts | >45 | >45 | >45 | - | 0.901 | - | / | Consistent Negative | RJ |
| P258 | Nasopharyngeal swab | Fever | Upper respiratory infection | >45 | >45 | >45 | - | 0.950 | - | / | Consistent Negative | RJ |
| P259 | Nasopharyngeal swab | Asymptomatic | Close contacts | >45 | >45 | >45 | - | 1.027 | - | / | Consistent Negative | RJ |
| P260 | Nasopharyngeal swab | Cough | Upper respiratory infection | >45 | >45 | >45 | - | 1.066 | - | / | Consistent Negative | RJ |
| P261 | Nasopharyngeal swab | Fever | Upper respiratory infection | >45 | >45 | >45 | - | 0.911 | - | / | Consistent Negative | RJ |
| P262 | Nasopharyngeal swab | Asymptomatic | Close contacts | >45 | >45 | >45 | - | 0.936 | - | / | Consistent Negative | RJ |
| P263 | Nasopharyngeal swab | Asymptomatic | Close contacts | >45 | >45 | >45 | - | 0.921 | - | / | Consistent Negative | RJ |
| P264 | Nasopharyngeal swab | Cough | Upper respiratory infection | >45 | >45 | >45 | - | 1.066 | - | / | Consistent Negative | RJ |
| P265 | Nasopharyngeal swab | Asymptomatic | Close contacts | >45 | >45 | >45 | - | 0.901 | - | / | Consistent Negative | RJ |
| P266 | Nasopharyngeal swab | Fever | Upper respiratory infection | >45 | >45 | >45 | - | 0.936 | - | / | Consistent Negative | RJ |
| P267 | Nasopharyngeal swab | Asymptomatic | Close contacts | 43.65 | >45 | >45 | - | 0.936 | - | / | Consistent Negative | RJ |
| P268 | Nasopharyngeal swab | Cough | Upper respiratory infection | >45 | >45 | >45 | - | 1.027 | - | / | Consistent Negative | RJ |
| P269 | Nasopharyngeal swab | Fever | Upper respiratory infection | >45 | >45 | >45 | - | 0.921 | - | / | Consistent Negative | RJ |
| P270 | Nasopharyngeal swab | Cough | Upper respiratory infection | >45 | >45 | >45 | - | 1.066 | - | / | Consistent Negative | RJ |
| P271 | Nasopharyngeal swab | Asymptomatic | Close contacts | >45 | >45 | >45 | - | 1.027 | - | / | Consistent Negative | RJ |
| P272 | Nasopharyngeal swab | Dyspnea | Pneumonia | >45 | >45 | >45 | - | 0.904 | - | / | Consistent Negative | RJ |
| P273 | Nasopharyngeal swab | Cough | Upper respiratory infection | >45 | >45 | >45 | - | 1.082 | - | / | Consistent Negative | RJ |
| P274 | Nasopharyngeal swab | Fever | Diabetes | >45 | >45 | >45 | - | 0.936 | - | / | Consistent Negative | RJ |
| P275 | Nasopharyngeal swab | Asymptomatic | Close contacts | >45 | >45 | >45 | - | 1.066 | - | / | Consistent Negative | RJ |
| P276 | Nasopharyngeal swab | Asymptomatic | Close contacts | >45 | >45 | >45 | - | 1.011 | - | / | Consistent Negative | RJ |
| P277 | Nasopharyngeal swab | Asymptomatic | Close contacts | >45 | >45 | >45 | - | 0.877 | - | / | Consistent Negative | RJ |
| P278-1 | Nasopharyngeal swab | Asymptomatic | Close contacts | N/A | 36.25 | N/A | + | 1.068 | - | - | False Positive | RJ |
| P278-2 | Nasopharyngeal swab | Asymptomatic | Close contacts | >45 | >45 | >45 | - | 1.068 | - | / | Consistent Negative | RJ |
| P279-1 | Nasopharyngeal swab | Asymptomatic | Close contacts | N/A | 36.89 | N/A | + | 1.030 | - | - | False Positive | RJ |
| P279-2 | Nasopharyngeal swab | Asymptomatic | Close contacts | >45 | >45 | >45 | - | 1.030 | - | / | Consistent Negative | RJ |
| P280 | Oropharyngeal swab | Fever, Cough, Headache | COVID-19 suspected | N/A | >40 | 34.27 | Uncertain | 1.172 | + | / | Uncertain Positive | JNCDC |
| P281 | Fecal | Fever, Cough, Rhinobyon | Close contacts | N/A | 30.20 | 30.46 | + | 1.453 | + | / | Consistent Positive | JNCDC |
| P282 | Fecal | Fever, Cough, Rhinobyon | COVID-19 suspected | N/A | 38.50 | >40 | - | 1.250 | + | / | False Negative | JNCDC |

Note: N/A, Detection of this gene that was not supported by the kit. “+”, Positive; “-”, negative; /, PCR product was not analyzed by NGS. Samples in aquamarine background stood for SENA negative but rRT-PCR positive or uncertain diagnosis. Samples in red background stood for SENA positive but rRT-PCR negative or uncertain diagnosis.

